# The burden, clinical outcomes, and risk factors related to neglected tropical diseases and malaria in migrant populations in the Middle East and North Africa: A systematic review and meta-analyses

**DOI:** 10.1101/2025.08.16.25333818

**Authors:** Eman Elafef, Taha Maatoug, Stella Evangelidou, Helena Martí Soler, Asad Adam, Ahmed Hamed Arisha, Mahmoud Hilali, Sally Hargreaves, Ibrahim Bani, Farah Seedat, Ana Requena-Méndez

**Affiliations:** Blue Nile National Institute for Communicable Diseases, University of Gezira, Sudan; Barcelona Institute for Global Health (IS Global-University of Barcelona), Spain; School of Health & Medical Sciences, City St George’s University of London, London, UK; Université de Sousse, Tunisia; Faculty of Veterinary Medicine, Badar University in Cairo, Egypt; Chronic Disease Epidemiology Department, Yale School of Public Health, USA; Department of Infectious Diseases, Karolinska University Hospital, Stockholm, Sweden; CIBERINFEC, ISCIII, Centro de Investigación Biomédica en Red de Enfermedades Infecciosas, Barcelona, Spain

**Keywords:** Migrants, Malaria, Neglected tropical diseases, Epidemiology, MENA, Meta-analysis

## Abstract

**Introduction:** This systematic review investigates the burden, clinical outcomes, and risk factors of neglected tropical diseases (NTDs) and malaria in the Middle East and North African region (MENA), highlighting the urgency and scope of these health challenges.

**Methods:** We searched six databases for peer-reviewed literature and additional sources to capture grey literature in any language from 2000 to 28 August 2024. Studies were included if they provided primary data on outcomes in migrants. Primary outcomes were prevalence, incidence, and mortality. Peer-reviewed articles were critically appraised using JBI tools, while the AACODS checklist was used for grey literature. Estimates were pooled using random-effects meta-analysis where possible or synthesised narratively. (PROSPERO-CRD42023401694)

**Results:** We included 39 studies with 81,678 migrants across 11 countries for NTDs and 16 studies encompassing 12,823 migrants across five countries for malaria. The pooled prevalence of specific NTDs among migrants was 4.7% for hookworm (95% confidence interval [CI] 0.9-11.3, I²=99%), 1.8% for *Trichuris trichiura* (0.3-4.3, I²=98%), 1.75% for *Ascaris lumbricoides* (0.6-3.5, I²=96%), and 1.8% for taeniasis (0.3-4.4, I²=98%). Compared with non-migrants, migrants exhibited higher prevalence rates for hookworm (1.8% vs. 0.03%), *Ascaris lumbricoides* (0.3% vs. 0%), *Trichuris trichiura* (0.5% vs. 0%), dengue (26% vs. 3.5%), and chikungunya (4.2% vs. 0.5%). Migrants had a higher proportion of confirmed cases of schistosomiasis (0.21%-20.3% vs 0-0.013%), cystic echinococcosis (7.4% vs 3.5%), and dengue (57.2% vs 56.4%) among suspected cases compared to non-migrants. Case fatality rates were 3.1% for dengue and 0.2%-1.5% for malaria. Malaria incidence was only reported in Sudan (Internally displaced persons: 6.8/1,000; refugees: 2.72/1,000; refugees <5 years old: 7.3/1,000). While hospitalisation and ICU rates for malaria were 25.8% and 1.3%, respectively, severe malaria was higher in non-migrants compared to migrants in Qatar (50% versus 5.2%, respectively).

**Conclusions:** Despite a wide range of diseases reported in 55 studies, there were gaps in the evidence, primarily related to risk factors, clinical outcomes, and the sub-region of North Africa. We generally found that migrants were disproportionately affected by both NTDs and malaria, especially in the Middle East.

**Key questions:** *What is already known?:*

- The Middle East and North African (MENA) region has experienced a rise in migration from neglected tropical diseases (NTDs)- and malaria-endemic countries.
- Without adequate access to prevention and treatment, there is a risk of reintroducing or increasing the burden of NTDs and malaria in MENA countries that have achieved or made progress towards their elimination.
- Despite the critical importance of understanding the burden, clinical outcomes, and risk factors of NTDs and malaria among migrants in the MENA region, little is known about these diseases in this population.
- Such knowledge is essential for designing effective interventions, delivering targeted care, and monitoring morbidity and mortality.

*What are the new findings?:*

- Overall, migrants in the MENA region were disproportionately affected by NTDs and malaria.
- They had a higher prevalence of soil-transmitted helminths and a higher seroprevalence of dengue and chikungunya than non-migrants.
- There was also a higher proportion of confirmed cases of schistosomiasis, cysticercosis, echinococcosis, dengue, scabies, and malaria among suspected cases.
- Migrants also represented a higher percentage of reported cases of malaria, dengue, scabies, leprosy, schistosomiasis, and CL.
- Outcome and risk factor data and North African data on NTDs and malaria were limited.

*What do the new findings imply?:*

- Migrants in most MENA countries are disproportionally affected by NTDs and malaria, highlighting their vulnerability as at-risk groups.
- However, their needs have not been prioritised in research. There is an urgent need to strengthen research on NTDs and malaria for migrants in the region to fill existing knowledge gaps and guide evidence-based approaches for effective prevention, early diagnosis, and treatment of NTDs and malaria to serve migrant populations better.

## INTRODUCTION

The Middle East and North African (MENA) region serves as a point of origin, transit, and destination for different migrant groups and is marked by significant health disparities, ongoing regional and sub-regional conflicts, and complicated mixed-migration flows.[1] In 2020, the MENA region hosted 42.7 million international migrants, making it the third largest region receiving international migrants, including forced migrants, labour migrants, refugees, asylum seekers, and internally displaced persons (IDPs).[2] This trend has only intensified with the mass displacement of over 1.7 million people in Gaza and the armed conflict in Sudan, which has forced 2.1 million people to cross borders and displaced 7.9 million within the country.[3]

Globally, migrants face heightened risks of neglected tropical diseases (NTDs) and malaria due to a complex interaction of several factors, including higher disease prevalence in their countries of origin and socio-economic factors, living conditions, and barriers to healthcare access in their countries of transit and destination.[4] NTDs are a group of 23 diseases, such as trachoma and scabies,[5] and are related to environmental and socioeconomic conditions in low-income communities (i.e., lack of appropriate medical infrastructure or sanitation).[6] Migrants can experience precarious living and working conditions throughout their migration journey, which can make them susceptible to NTDs.[6] McCarthy et al. (2013) found that schistosomiasis was diagnosed in 13% of 2,804 migrants from 153 countries at 41 GeoSentinel clinics across 19 countries,[7] which is seven times higher than global estimates.[8] Similarly, understanding the movement patterns of migrants and the factors that increase their risk of malaria is essential to improve migrant health outcomes, ultimately protecting the health of migrants and host communities.[9] Indeed, the World Health Organization’s 2030 roadmap for NTDs, and the global strategy for malaria, recognises migration as a risk factor and migrants as a marginalised high-risk group, calling for intervention strategies to ensure timely and free access to prevention, diagnosis, and treatment for migrant populations.[10–12]

Despite this, little is known about the burden, clinical outcomes, or risk factors of NTDs and malaria in migrants living in the MENA region. This knowledge gap is of critical importance as many countries in the region, most recently Egypt, have achieved malaria elimination,[13] and increased migration from endemic countries could increase the risk of re-introduction if effective prevention interventions are not in place.[13] Likewise, many migrants in the region live in poor conditions, which increases their vulnerability to NTDs,[14] and clinicians may be unfamiliar with these diseases, leading to delays in diagnosis and treatment.[15][10,11]This systematic review aimed to identify, appraise, and synthesise the empirical evidence on the burden, clinical outcomes, and risk factors related to NTDs and malaria in migrant populations in the MENA region.

## METHODS

This systematic review and meta-analysis are reported according to PRISMA 2020 guidelines.[16] The protocol is registered on PROSPERO (CRD42023407748) and published.[17]

### Search strategy

We searched Medline, Embase, Web of Science, Cumulative Index to Nursing and Allied-Health Literature (CINHAL), Index Medicus for Eastern Mediterranean Region and Q Science for articles published between 2000 to August 2024 in any language, combining three sets of search terms using free text and subject heading for migrants, NTDS and malaria, and countries in the MENA region (Annex 1). We also conducted an extensive search of grey literature sources from the following websites: World Health Organization, International Organization for Migration, United Nations High Commissioner for Refugees, United Nations Department of Economic and Social Affairs, Médecins Sans Frontières, Commission of Refugee (COR), and Ministry of Health websites for each country in the MENA region, reference-checked all included studies and relevant systematic reviews, and contacted experts for additional references.

### Eligibility criteria

We included studies that met the following criteria:

- Population: Migrants, defined as people who move away from their usual residence, whether within a country or across an international border, temporarily or permanently, and for various reasons.[19] This included refugees, asylum seekers, undocumented migrants, labour migrants, and IDPs.[20]
- Geographical scope: 18 MENA countries (Algeria, Bahrain, Egypt, Iraq, Jordan, Kuwait, Lebanon, Libya, Morocco, Oman, Palestine, Qatar, Saudi Arabia, Sudan, Syrian Arab Republic, Tunisia, United Arab Emirates (UAE), and Yemen).[18]
- Health outcomes: Primary data on the burden (e.g., prevalence, incidence), clinical outcomes (e.g., treatment results, long-term morbidity, mortality) or risk factors of NTDs and malaria in migrants.
- Diseases of interest: Malaria and the following NTDs: buruli ulcer, dengue and chikungunya, dracunculiasis, echinococcosis, human African trypanosomiasis, leishmaniasis, leprosy, lymphatic filariasis, mycetoma, onchocerciasis, rabies, scabies and other ectoparasites, schistosomiasis, soil-transmitted helminthiases, snakebite envenoming, taeniasis/cysticercosis, trachoma, and yaws.[5]
- Study design: Cohort studies, cross-sectional studies, case-control studies, systematic reviews, narrative reviews, and clinical trial studies (data only extracted from baseline characteristics or the control group as appropriate).
- Time frame and language: Published in any language from 2000 to August 2024.

We excluded the following NTDs because they are not endemic in the countries from which migrants living in the MENA region originate: foodborne trematodes, chromoblastomycosis, other deep mycoses, and Chagas disease. We excluded studies if they did not meet the key definitions for migrants, MENA countries, or NTDs or malaria; studies published before 2000; and studies in which migrant data could not be disaggregated from host populations. We excluded case series, case reports, qualitative studies, and studies not providing primary data. Systematic and narrative reviews were not included but were comprehensively reviewed for additional studies providing primary data.

Records were imported into Rayyan,[21] and deduplicated. Two reviewers (EE & TM) independently screened the titles, abstracts, and full texts of all identified records, and disagreements were discussed with a third reviewer (FS) when necessary.

### Data extraction

EE and TM independently extracted relevant data using an a priori-defined extraction sheet, which was piloted and refined before implementation. Extracted data were cross-checked for consistency, and any disagreements were resolved through discussion with a third reviewer (FS) to ensure accuracy and reliability. Data extracted included study information (country, publication year, design, and study setting), study population (sample size, migrant and host population definitions and characteristics), disease(s), outcome definitions, and results (burden, clinical outcomes, risk factors), (Annex 2). From studies with adjusted and unadjusted analyses, we prioritised the adjusted analysis, and we calculated any missing statistical parameters of importance and variability measures (e.g., 95% CIs), when data permitted. We contacted the authors of included studies to obtain missing or additional data.

### Quality appraisal

EE and TM independently appraised the risk of bias in studies using Joanna Briggs Institute checklists for peer-reviewed articles.[22] The checklists contain eight domains for cross-sectional studies, nine for prevalence and 10 for case-control studies, including participant and setting description, outcomes/exposure measurements, and appropriate statistical analyses. Every criterion was rated ‘yes’, ‘no’, ‘unclear’ or ‘not-applicable. The AACODS checklist was used to appraise grey literature. This checklist contains six domains: Authority, Accuracy, Coverage, Objectivity, Date, and Significance. Each domain is assigned a rating of ‘yes’, ‘no’, or ‘unclear’. Studies were not excluded based on quality, however, the results of quality assessments contributed to the analysis and the discussion.[23] Any disagreements were resolved by discussion, involving a third reviewer if necessary (FS).

### Data analysis

Meta-analyses were conducted only when there was sufficient data and homogeneity. A minimum of three data points was required to meet the volume threshold, and homogeneity was assessed based on population type, diseases, outcome measurements, and consistent outcome definitions. When these criteria were met, we conducted a random-effects meta-analysis on single proportions using the Metaprop function in R software (version 4.2.2).[24] We assessed heterogeneity among studies by inspecting the forest plots, using the chi-squared test for heterogeneity with a 10% level of statistical significance and using the I² statistic. Where data volume or homogeneity was insufficient, we synthesised findings narratively and presented them in tables, text, and figures. We calculated statistical parameters, (i.e., risk ratios), when authors had not provided them but adequate data were available.

## RESULTS

### Characteristics of the included studies

Our search identified 1,598 database records and 80 from additional sources. After assessing the titles, abstracts, and full texts, we included 55 peer-reviewed studies (Figure 1, Annex 3). Sixteen studies, including 12,823 migrants, reported on malaria,[25–40] and 39 studies, including 82,728 migrants reported on NTDs; specifically 12 on cutaneous leishmaniasis (CL);[41–52] eight on intestinal helminthic infection;[53–60] five on dengue and chikungunya;[61–65] three on schistosomiasis, cystic echinococcosis,[66–68] trachoma,[69–71] and scabies,[72–74]; two on leprosy[75,76] and cysticercosis [77,78] and one on lymphatic filariasis.[79] Studies spanned eleven countries, 80% (n=44) in the Middle East,[25–28,32–35,37,39–48,50–63,65,66,66,67,67,72–79] and 20% (n=11) in North Africa (Algeria, Morocco, Sudan [n=9]). Thirty-four studies investigated non-nationals, [25–28,32–35,37,39–43,51,53–59,61–63,65–68,75–79]ten refugees,[29,36,44–48,50,69,71] and 11 IDPs.[30,31,38,49,52,60,64,70,72–74]

**Figure 1.**
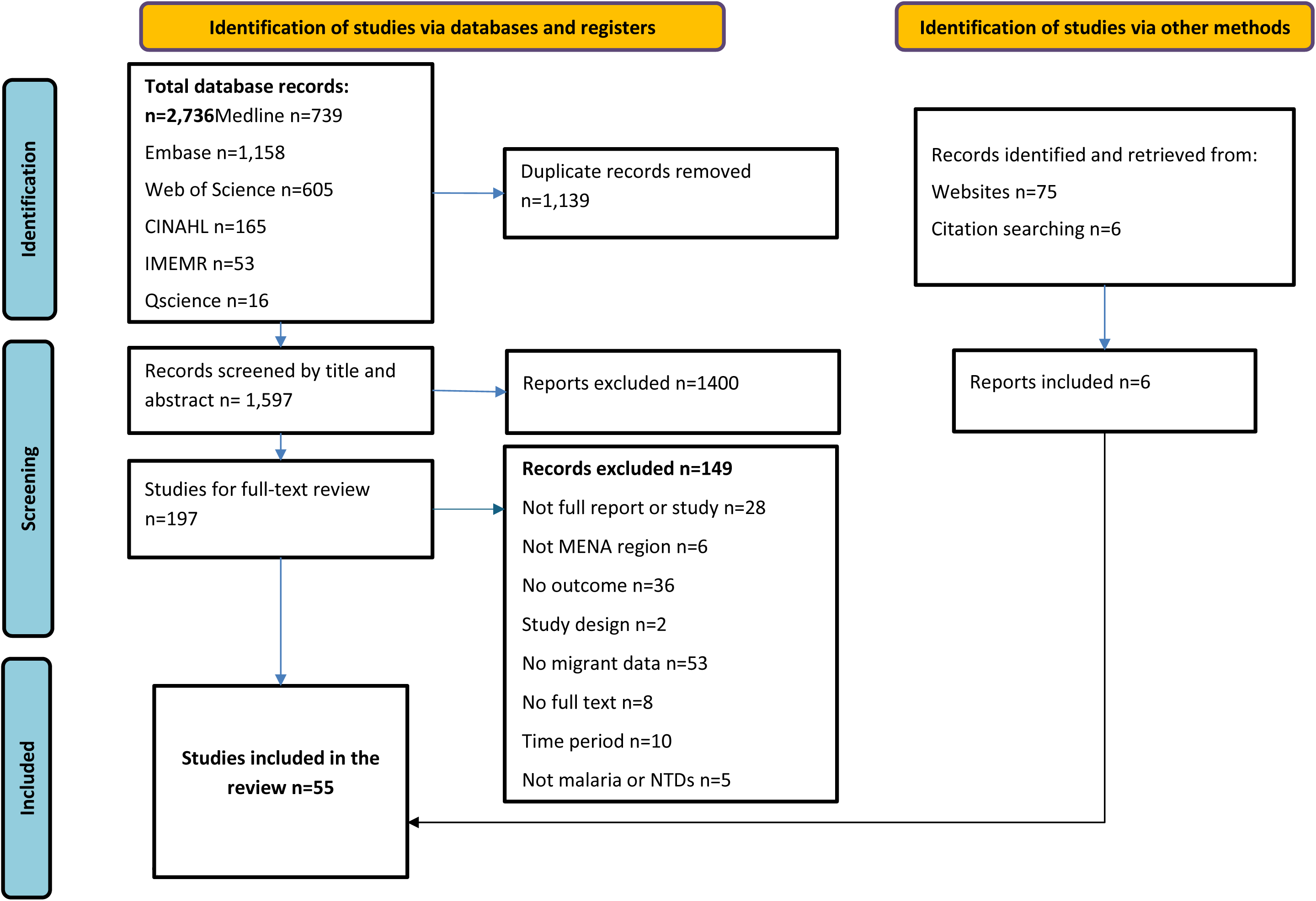
**Flowchart of included studies**

Fourteen studies reported prevalence or seroprevalence of diseases, [49,53–59,61,69–71,77,79] four examined incidence,[31,36,50,65] 13 documented number of confirmed cases among suspected cases, [25,29,30,35,38,48,60,62,64,66,67,73,74] and a majority (n=24) investigated the distribution of confirmed cases between migrants and non-migrants.[26–28,32–34,37,39–47,51,63,68,72,75,76,78] Only six studies were conducted on clinical outcomes, including severe malaria, severe dengue haemorrhagic fever, and the case fatality rate of dengue and malaria.[30,33,34,36,48,64] Finally, 14 studies reported risk factors for dengue and chikungunya,[61] cystic echinococcosis,[66] schistosomiasis,[66] malaria,[29,31,33–35,38] soil-transmitted helminths (STH),[53,54,56] CL,[52] and cysticercosis.[77] Thirty-four studies compared the burden of diseases in migrant versus non-migrant populations[26–29,32,34,35,37,39–44,46,47,51–55,61–63,65–68,72,73,75,76,78] and 21 were only conducted in a migrant group.[25,30,31,33,36,38,45,48–50,56–60,64,69–71,74,77,79]

### Quality assessment

None of the 40 cross-sectional studies were judged as having a low risk of bias across all domains. The highest risk of bias was observed in domains related to confounding factors, with 37 studies failing to apply any strategies to address potential confounding factors, and two studies not discussing variables that could influence the outcomes. Additionally, a high risk of bias was identified in outcome measurement in 10 studies, either because of sole reliance on clinical diagnosis or a lack of detailed information about the diagnostic method used.

Five of the 14 prevalence studies had a low risk of bias in all domains, particularly for domains related to the study subjects and setting, using an appropriate sample frame, sufficient study acceptance and completion rate, and using valid methods and standard criteria to measure the condition. The domains at unclear risk of bias included sample size adequacy and appropriateness of sampling method, as the study did not provide details in these areas (Figure 2).

**Figure 2.**
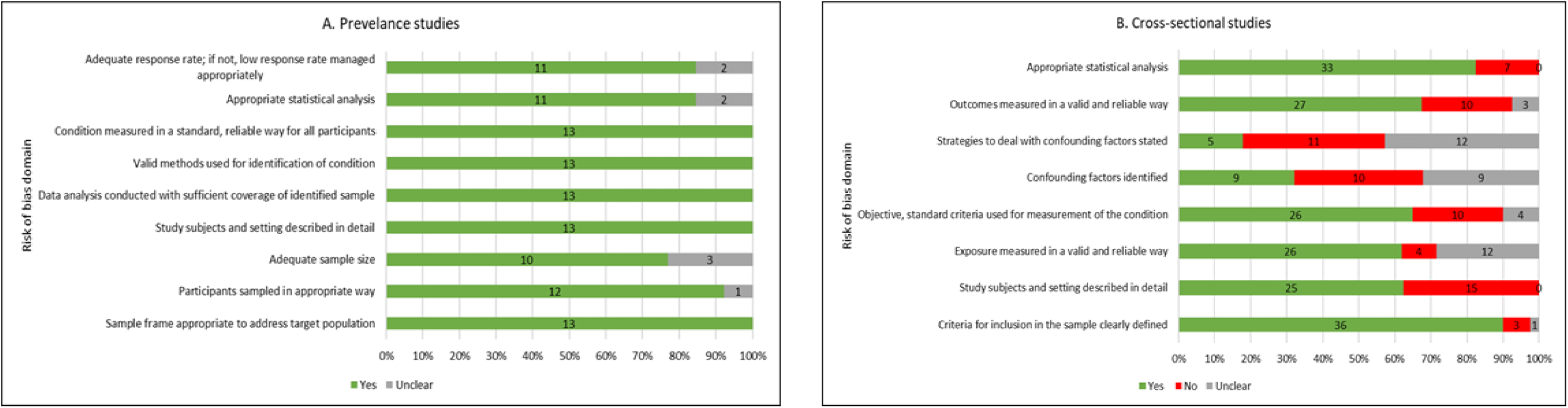
Risk of bias across 55 peer-reviewed studies using JBI critical appraisal tools for A) Prevalence and B) Cross-sectional studies. Notes: Five of the 14 prevalence studies had a low risk of bias in all domains. The two JBI domains on confounding factors were not applicable for 12 of 40 cross-sectional studies.

### Burden of disease

#### Soil-transmitted helminths

Seven studies evaluated the prevalence of STH,[53–59]. After pooling the data from four screening studies, including 25,491 adult migrants in Gulf Cooperation Council (GCC) countries,[56–59] we found that, based on microscopic stool examination, the prevalence of hookworm was 4.7% (0.9-11.1, n=259,I²=99%), *Trichuris trichiura* was 1.8% (0.4-4·3, n=125,I²=98%), and *Ascaris lumbricoides* was 1.7% (0.6-3.5, n=166, I²= 96%),(Figure 3). The other three studies, also based on microscopic stool examination, compared the prevalence of STH amongst long-term migrants (n=19,929) vs non-migrants (n=9,357) attending hospitals from 2005-2014 in Qatar. They found no cases of *Ascaris lumbricoides* and *Trichuris trichiura* among non-migrants, while in migrants, prevalence was 0.3% for *Ascaris lumbricoides* (n=62) and 0.5% for *Trichuris trichiura* (n=97). Additionally, migrants exhibited a higher prevalence of hookworm at 1.8% compared to 0.03% in non-migrants (unadjusted risk ratio (RR) 55.56 [17.8-173.04]).[53–55].

**Figure 3.**
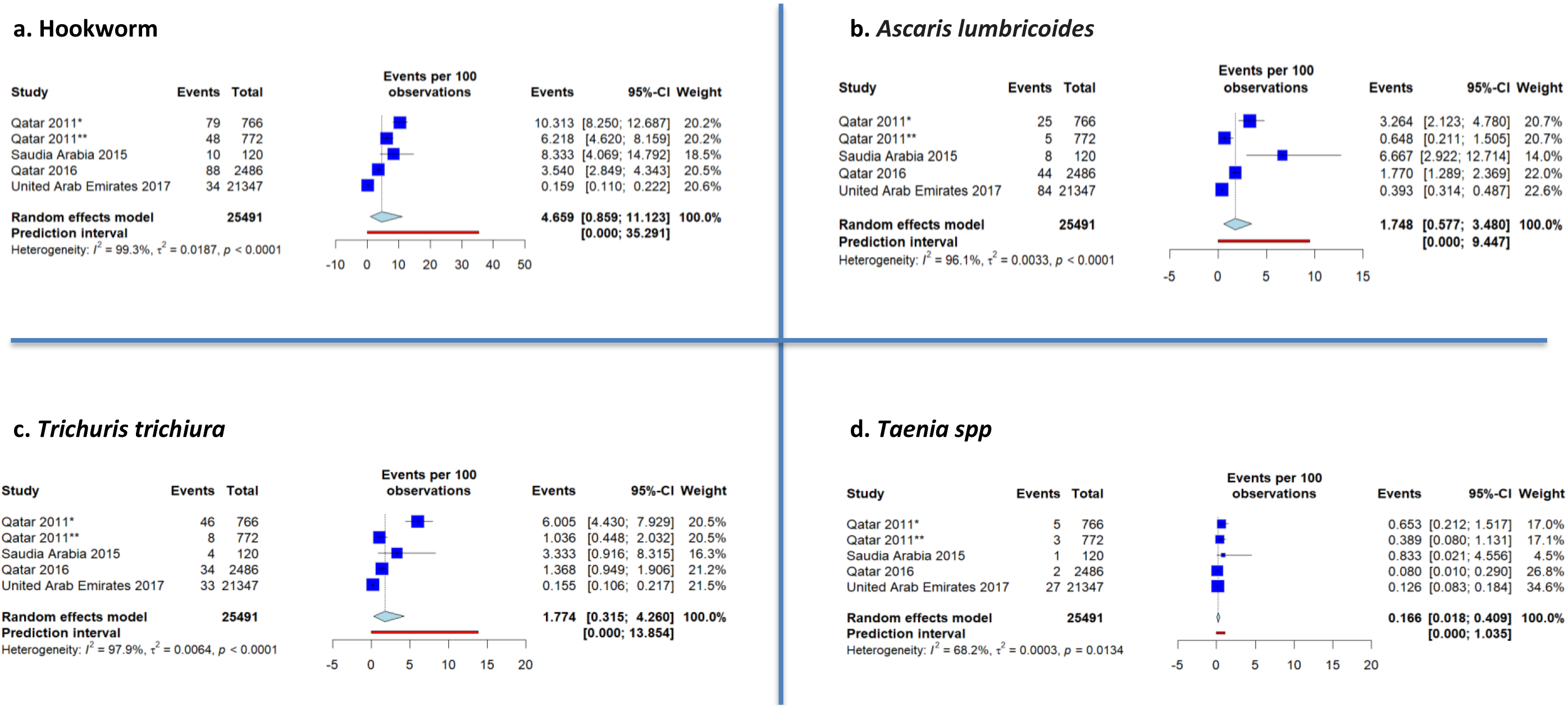
**Forest plots of the pooled prevalence of intestinal helminths among migrants in the Middle East and North African region: A) Hookworm, B) *Ascaris lumbricoides*, C) Trichuris, and D) Taenia [55–58]** Note: *Newly arrived migrants **Long-term resident migrants

There were only two studies on risk factors for STH from Qatar. Abu-Madi et al. (2010) reported that, compared with other nationalities, Asian migrants had a higher prevalence of hookworm (3.38% vs 0%, p<0.001), *Ascaris lumbricoides* (0.58% vs 0%, p<0.001), and *Trichuris trichiura* (0.91% vs 0%, p<0.001).[53] (see Annex 4). In 2016, Abu-Madi et al. found that hookworm infections were more common among newly arrived migrants in Qatar who were aged 38-58 years (p=0.011) and male (4.4% [3.3-5.8] vs 2.8% [2.0-3.9], p=0.033). Additionally, parasitic load (mean eggs per gram of faeces) was higher amongst those aged 38-58 years (p=0.004), and females had a higher mean egg count (2.5, SD = 0.8 versus 2.2, SD = 0.4, p=0.032).[56]

#### Taeniasis, cysticercosis, and other helminthic infections

The pooled prevalence of taeniasis in four studies containing 25,491 adult migrants in GCC countries, based on microscopic stool examination, was 0.2% (0.0-0.4, n=38, I²=68%).[56–59] A study in Kuwait found a seroprevalence of cysticercosis of 4.2% in a sample of 1,000 newly arrived migrants.[77] A study in Iraq found 2 (0.05%) stool positive cases among 3,925 suspected taeniasis in IDPs. [60] Another study in Qatar compared the distribution of neurocysticercosis cases by migration status, using radiographic imaging, histology, and electrocardiogram, and found that migrants represented a higher percentage (98.8% n=415/420) than non-migrants [78] Another study in Kuwait reported a higher proportion of cystic echinococcosis (positive serology) in migrants among suspected cases (7.4% [n=42/568] vs 3.5% [n=15/423], unadjusted RR 2.08 [1.2-3.7]).[66] In this study, Bedouin (3.7%,n=21/568), Syrians (1.7%, n=10/568), and North Africans (0.9%,n=5/568) had a higher proportion of cystic echinococcosis compared with other nationalities (p<0.001).[66] Another study in migrant workers in Kuwait found a prevalence of filarial infection of 18.3% (192/1050) using immunochromatographic test (ICT) methods and 20.3% (n=213) with the TropBio assay. Additionally, microscopy detected 32 cases (3%) of *Wuchereria bancrofti*, with a mean microfilaria count of 816 per millilitre.[79]

#### Schistosomiasis

Two studies investigated the prevalence of schistosomiasis among migrants, finding a prevalence of 0.3% among 772 long-term resident migrants in Qatar[57] and 0.83% among 120 migrant workers in Saudi Arabia, determined by microscopic stool examination[59]. Two studies compared the proportion of positive cases among suspected schistosomiasis cases between migrants and nationals. In Saudi Arabia, migrants exhibited a higher positive rate (0.2%, n=4/1891) than nationals (0.01%, n=29/232,364),(unadjusted RR 16.9[6.0-48.2]) based on stool microscopy.[67] In Kuwait, migrants with suspected schistosomiasis had a rate of 20.3% (n=186/916), whereas in non-migrants there were zero positive cases, based on serological screening.[66] In this study, being Egyptian (14.1%, n=129/916) and Bedouin (6.1%, n=56/916) was associated with schistosomiasis (p<0.001).[66] A third study in Morocco reported that all cases (n=27) of schistosomiasis were in migrants.[68]

#### Cutaneous leishmaniasis

A study of CL among IDPs in Sudan found the prevalence of CL is 55.7% (n=688/1236), using tissue smear microscopy.[49] Another study on the incidence of CL in Syrian refugees residing in Lebanon, found a rate of 0.21% in 2013 and 0.04% in 2014 (1,683 cases from 2013-2019).[50] Another three studies also investigated CL among Syrian refugees; one found that the proportion of positive among suspected CL cases in Syrian refugees in Lebanon was 11.6% (n=110/948), based on microscopy and PCR.[48] The other two studies reported that 92.1% of clinically diagnosed CL cases among 558 Syrian refugees in Jordan were imported. In contrast, 2.5% were locally acquired.[44,45]

Seven studies compared the distribution of CL among migrants and non-migrants, with conflicting results (see Annex 5). Three studies based on laboratory analysis showed that migrants represented a higher percentage of CL cases compared with non-migrants; 96.5% (n=1206/1256) of CL cases in Lebanon were Syrian refugees,[47] and 54% (n=5393/9962) and 80.5% (n=132/164) in other two studies in Saudi Arabia were migrants. [41,42] [42][41]By contrast, four studies found that non-migrants represented a higher percentage of CL; based on clinical diagnosis migrants represented 28.6% of CL cases in Iraq[46] and 44.97% (n=559/1243) in Jordan[44] and based on microscopic examination, 9.3% (n=559/1234) in Asir and 46.9% (n=739/1575) in Tabuk in Saudi Arabia were migrants.[43,51]

Only one study on treatment outcomes described an initial cure rate of CL in migrants in Lebanon of 81.8%.[48] Another study on risk factors for CL, found that a history of displacement was associated with CL (unadjusted OR 5.18 [3.84-6.98]).[52]

#### Dengue and chikungunya

Five studies compared the burden of dengue between migrants and non-migrants, finding that migrants have a higher burden.[61–65] In Jizan, Saudi Arabia, although migrants represented a lower percentage of confirmed dengue cases (26.6%, n=910/3427), diagnosed by PCR or serology from 2015 to 2020, the incidence rate from 2015 to 2019 surpassed that of non-migrants, except in 2016. This disparity was particularly pronounced in 2015 when the incidence for migrants was 26.8/100,000 compared to 14.5/100,000 among non-migrants.[65] Likewise, in Qatar, the seroprevalence of dengue among blood donors was six times higher among migrants (26% [n=466/1792]) than among nationals (3.5% [n=7/200]) (unadjusted RR 5.9 [2.8-12.3]).[61] Another study in Qatar found that migrants (98%, n=163) represented more of the serologically confirmed dengue cases than nationals.[63] By contrast, only one study in Saudi Arabia found that migrants had a similar rate of serologically confirmed cases among suspected dengue cases as nationals (57.2%,n=1752 vs. 56.4%,n=4885).[62] The study investigating blood donors in Qatar also compared the seroprevalence of chikungunya but found a non-significant higher rate in migrants compared to nationals (4.2% (n=75/1792) vs 3.5%, (n=7/200), unadjusted RR 1.14 [0.5-2.5]).[61]

There was also only one study on clinical outcomes, which found that 15.7% of 204 Sudanese IDPs had dengue, 75% of whom had severe disease with haemorrhagic fever, and 3.1% died.[64] Only one study examined risk factors for dengue and chikungunya, finding that Asian migrants had higher odds of being seropositive for anti-DENV and anti-CHIKV antibodies than those from Qatar (aOR 0.23 [0.10– 0.53]), the Middle East (aOR 0.09 [0.05–0.16]), and North Africa (aOR 0.37 [0.20–0.67]), even after adjusting for age and other arboviruses.[61]

#### Other parasitic neglected tropical diseases

Three studies investigated ectoparasites among IDPs in Iraq, two of which compared the burden between migrants and non-migrants.[72–74] In one study, migrants accounted for 95% of scabies cases (n=145/195), a significantly higher percentage than non-migrants based on clinical criteria for scabies diagnosis.[72] Another study compared the proportion of positive cases among suspected scabies in migrants and nationals and found no difference (5.45% [n=342/6,264] versus 5.5%, [n=1612/29,183] respectively, unadjusted RR 0.99 [0.88-1.1]), based on clinical features, microscopy, or dermatoscopy.[73] A third study found that 45% of suspected scabies cases among Iraqi IDPs were microscopically confirmed (n=585/1300).[74]

#### Bacterial neglected tropical diseases

Three studies investigated the prevalence of trachoma, and two investigated the distribution of leprosy cases by migrant status.[69–71,75,76] The pooled prevalence of acute trachoma infection follicular among refugee children (n=3951) across three studies was 7.25% (2.8-13.6, I²=98%), based on clinical examination.[69–71] (Figure 4). The prevalence of trachoma trichiasis among adults refugee ranged from 0.75% (16/2139) to 3.07% (53/1734) based on clinical examination.[69,70] Studies on leprosy found that migrants were more highly represented than nationals among all cases in Saudi Arabia (57.4%,n=139/242)[76] and Kuwait (89.1%,n=41/46), based on WHO criteria and classification.[75]

**Figure 4.**
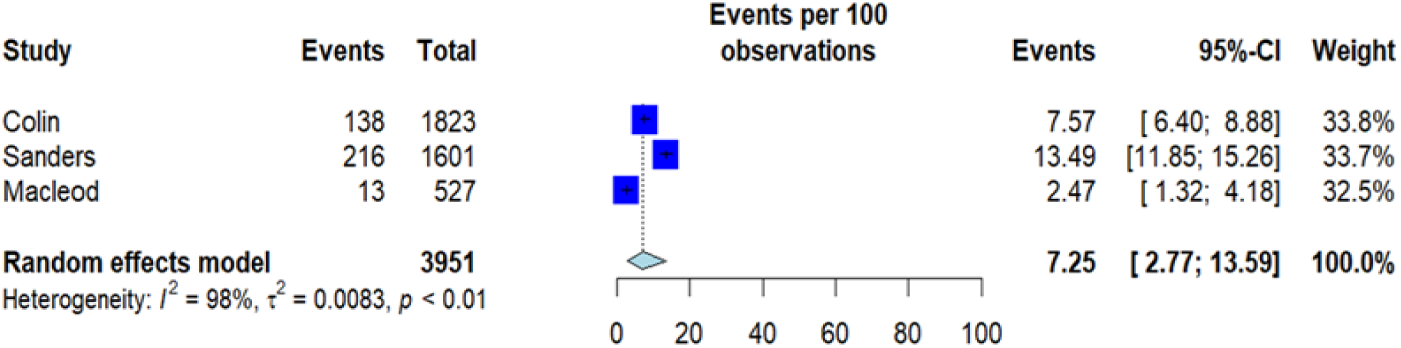
**Forest plot of the pooled prevalence of trachoma among migrant children in the Middle East and North African region**

#### Malaria

There were 16 studies on malaria; two reported malaria incidence,[31,36] five reported the proportion of positive among suspected malaria cases,[25,29,30,35,38] and nine reported the distribution of malaria cases by migration status.[26–28,32–34,37,39,40] The incidence of malaria was 6.8/1,000 among IDPs,[31] 2.72/1,000 among refugees and 7.3/1,000 among refugee children (<5 years) in Sudan.[36] Two studies compared the proportion of positive cases among suspected malaria cases between migrants and non-migrants. In Saudi Arabia, there was a higher rate in migrants (14.2%; n=19/134) versus non-migrants (7.8% n=11/141), based on polymerase chain reaction (PCR)[35]. In Sudan, there was a lower rate in migrants (6.6% [n=23/348]) versus non-migrants (17.9% [n=157/878]) based on microscopy.[29] Three studies investigated malaria cases in migrants alone (n=1,248). They found the following rates of positive cases: 8% (n=24/300) in IDPs in Khartoum, based on microscopy,[30] 61.4% (n=226/365) in IDPs in Darfur, Sudan, based on nested-PCR[38] and 76.4% (n=448/583) in non-nationals in Qatar based on microscopy,(Figure 5).[25] Three studies reported *P vivax* cases, which ranged from 3.7% (5/134) in Saudi Arabia,[35] 5.9% (23/384) in Sudan,[29] to 20% (118/583) in Qatar.[25]

**Figure 5.**
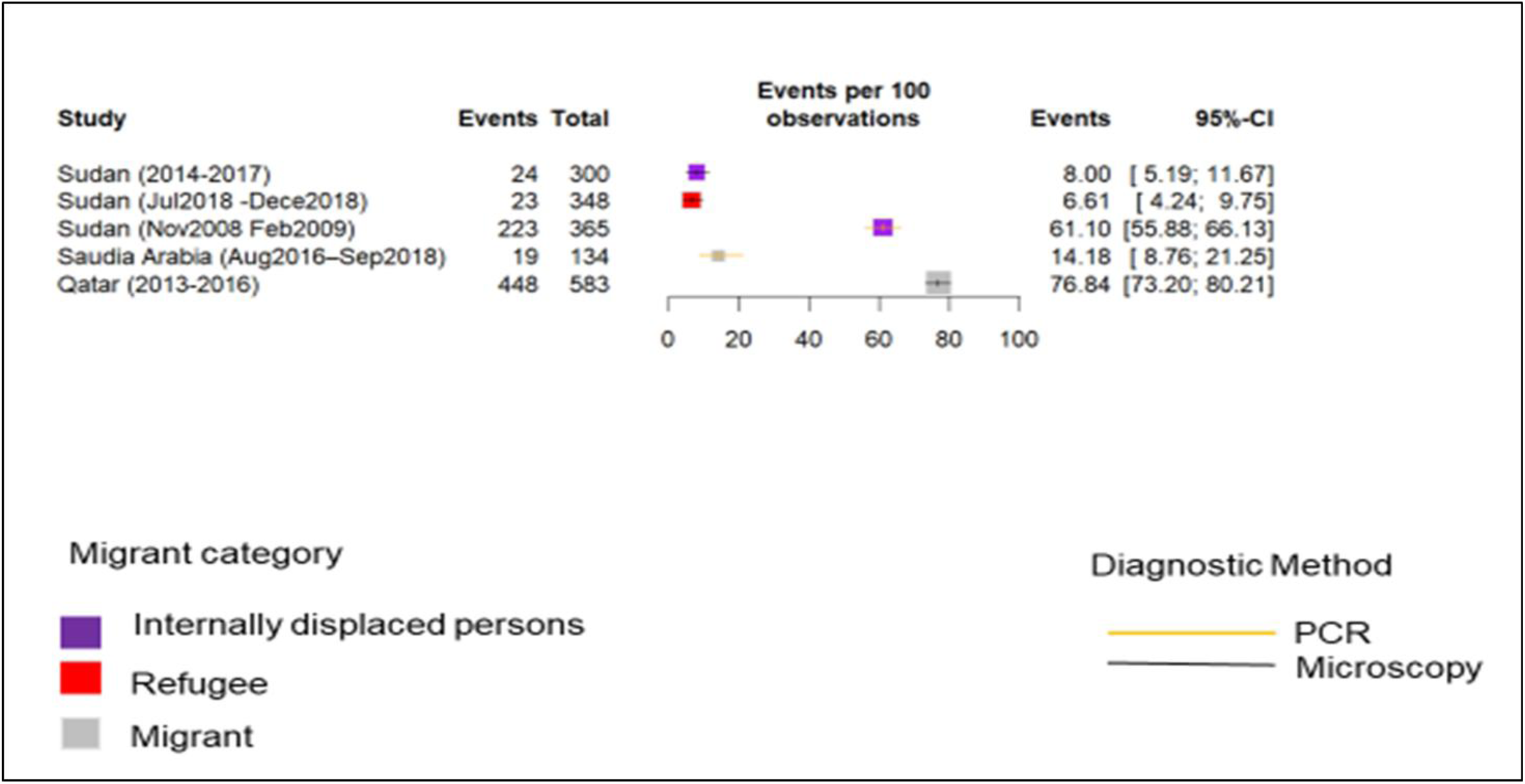
**Distribution of individual studies on the proportion of positive malaria cases among suspected cases in the MENA region** Abbreviations: PCR: Polymerase Chain Reaction

Ten out of 11 studies reported that migrants represented a higher percentage of malaria cases based on microscopy compared with non-migrants (Annex 6). The highest difference was in the UAE, where all cases were non-national (100%, n=629),[33] followed by Qatar (96.7%,n=4078/4092),[32] while the lowest difference was in Saudi Arabia (63%, n=19/30), diagnosed by PCR.[35] In contrast, the study in Sudan reported that non-migrants represented a higher proportion of malaria cases than Ethiopian refugees (migrants 12.8%, n=23/180, p<0.000).[29] Interestingly, the study in the UAE found that the median time from arrival to diagnosis in migrants was 2 days (interquartile range 1-4 days) and from symptoms onset to diagnosis was 5 days (3-8 days).

Six studies on risk factors found that water source, tribe, language, and percentage of income spent on food expenditure (as a proxy for socioeconomic status) were associated with malaria in migrants; however, water storage at home, educational level, occupation, marital status, species, housing condition, being a camp visitor or resident, knowledge, attitude, and practice were not associated (Annex 4).[29,31,33–35,38] In the adjusted analyses of a study in Sudan, the malaria attack rate within the last year was five times higher in IDPs who sourced water from a venting cart compared with a hand pump (OR 4.67[2.81-7.76]).[31] Additionally, spending 50% (OR 2.11,[1.08-4.11]) of all income (OR 1.97[1.09-3.61]) on food, speaking Dinka (a local language) compared with Arabic (OR 2.71,[1.43– 5.13]) and being from a Southern tribe compared with others (OR 1.94,[1.14-3.29]) resulted in higher odds of malaria in the last 12 months.

Four studies examined the clinical outcomes of malaria;[30,33,34,36] severe malaria was developed in 1.6% (5/300) of IDPs in Sudan,[30] and 0.3% (12/4,092) of non-nationals in the UAE.[33] Conversely, in Qatar, 50% [n=2/4] of non-migrant cases developed severe malaria compared with 5.2% [4/77] of migrants (p=0·02).[34] A study in the UAE found that, out of 692 malaria cases, all of which were in migrants, 25.8% were hospitalised, and 1.3% were admitted to ICU. Two studies examined malaria mortality; in the study in the UAE, case fatality was 0.14% (n=1)[33] while in Sudan, it was 12.1% (9.7-14.9) in all refugees and 15% (10.6-20.6) in children.[36] The annual malaria mortality rate in Sudan was 0.9 (0.7-1.1) per 1,000 in all refugees, while in children it was 4.1/1,000 (2.9-5.6).[36] Concerning risk factors for clinical outcomes, the study in the UAE found that *P. falciparum* was associated with hospitalisation and ICU compared with other species, but was not associated with death (Annex 4)

## DISCUSSION

In this systematic review and meta-analysis, we presented the evidence on the burden, risk factors, and clinical outcomes of NTDs and malaria among migrants in the MENA region. Despite a wide range of diseases reported in 55 studies, evidence gaps remain, primarily related to risk factors, clinical outcomes, and the sub-region of North Africa. Nevertheless, we found that in some countries, especially in the Middle East, migrants were disproportionately affected by both malaria and NTDs. Migrants showed a higher prevalence of STH and a higher seroprevalence of dengue than non-migrants, with up to 22.5% difference for dengue between the two groups. Among suspected cases, there was also a higher proportion of positive cases of schistosomiasis, cysticercosis, echinococcosis, dengue, scabies, and malaria in migrants, the difference ranging from 0.8% for dengue to 20.3% for schistosomiasis. Finally, migrants accounted for a higher proportion of reported cases of malaria, dengue, scabies, leprosy, schistosomiasis, CL and STH compared to non-migrants; indeed migrants constituted 100% of STH cases in GCC countries. Among studies that only included migrants, malaria, scabies, and trachoma were highly prevalent among IDPS and refugees. Regarding the region of origin, Asian migrants had a higher STH prevalence and higher odds of being seropositive for anti-DENV and anti-CHIKV antibodies compared with migrant from African and Mediterranean regions. Those of Bedouin, Syrian, or North African origin had a higher proportion of cystic echinococcosis, and being Egyptian or Bedouin was associated with schistosomiasis.

Our findings highlight both consistencies and differences within the included studies and with data from other regions. Generally, most studies within the MENA region and globally find that the burden of NTDs and malaria disproportionately affects migrant populations.[80]A systematic review of parasitic and vector-borne NTDs among migrants in Europe[81] reported a STH prevalence (4%) comparable to the MENA region; however, it found lower prevalence of filariasis and dengue in migrants (15.69% and 21.09%) compared to our findings in the MENA region, and diseases like CL, rare in migrants in Europe (prevalence of 0.42%), were highly endemic among Sudanese IDPs (55.7%). Similarly, the proportion of positive cases among suspected scabies in Iraq IDPs (5.5% to 45%), exceeded pooled European estimates, attributable to overcrowded camp conditions that facilitate transmission. In contrast, schistosomiasis and taeniasis were more common in migrants in Europe (10.8% and 1.98%) compared with the MENA region, likely reflecting Europe’s broader geographic origin and population diversity.These observed variations in disease burden among migrant populations appear closely tied to both origin and destination contexts. First, within GCC countries, studies predominantly identified STH and taeniasis among labour migrants, a pattern reflecting the region’s reliance on foreign workers from South and South-East Asia, and its mandatory pre-employment screening programme. This contrasts with European data where STH primarily affected Sub-Saharan Africa (SSA) migrants,[82,84] likely due to migration flows.[85] Schistosomiasis risk showed particularly stark disparities: African migrants in our analysis were twice as likely to be infected compared to Bedouin populations and 129 times more likely than Iraqi migrants, mirroring European findings of elevated risk among SSA migrants.[86]

Second, conflict-affected areas focussed on CL and hygiene-related NTDs. The Syrian crisis drove significant CL outbreaks among refugees in Jordan and Lebanon,[87,88] while IDP camps in Sudan and Iraq reported high burdens of hygiene-related NTDs (trachoma: 5.5-45% scabies prevalence) likely due to overcrowding and deficiencies in water, sanitation, and hygiene services. CL transmission dynamics are particularly complex, with risk influenced by both migrant status and local factors like vector exposure and prevention awareness – explaining why some studies in Iraq[46] and Saudi Arabia[43] found higher CL rates among non-migrants. Similarly, dengue affected over 15% of IDPs in Sudan,[64] a pattern attributable to the breakdown of environmental controls and health systems in protracted conflict settings - known risk factors for dengue transmission.[87] Importantly, IDPs were also found to have a higher proportion of malaria-positive cases than refugees,[29,30,38] as well as a high risk of scabies and trachoma.[70] This implies several challenges –IDPs are not eligible for international aid until their home country requests it explicitly and are not as well organised as refugee camps, making access to health care complex and limited research exists on the health issues of IDPs compared to refugees, possibly due to their lack of international status.[88]

Third, malaria distribution also followed geographic and demographic context: GCC nations reported primarily imported cases (higher among migrants than travellers), while endemic Sudan showed greater burden in local populations. Regional parasite distribution varied predictably, with *P. vivax* dominating among Asian migrants and *P. falciparum* among Africans. Together, these regional and global differences in disease burden amongst migrant populations demonstrate how geopolitical context, migrants’ status, country of origin, and vulnerability, diagnostic variability, and health system priorities interact to shape disease burdens,[89] underscoring the need for surveillance systems that capture this complexity and for interventions tailored to specific migrant subgroups.

Our findings have potential public health implications. Priority should be given to high-risk groups, such as Syrian refugees vulnerable to CL in Lebanon and Jordan, or Asian migrant workers in the GCC at risk of STH, with interventions like pre-departure and post-arrival screening and deworming. Effective prevention and early diagnosis for migrants including inclusive care is essential to prevent the re-introduction of diseases like malaria.[90]Indeed, malaria outbreaks have occurred in MENA countries that achieved elimination; falciparum malaria was reintroduced in Oman after decades due to imported cases among refugees, leading to 21 local cases[91]Climatic conditions in the MENA region could increase mosquito proliferation and together with the influx of migrants from endemic areas could facilitate malaria transmission.[12]

Additionally, clinicians and administrative staff involved in screening and health-care provision should receive specific training, to deliver proficient health services for NTDs and malaria,[92] particularly in malaria-free countries. Surveillance systems should include migrants, IDPs, and refugees indicators in health information systems to evaluate the burden of malaria and NTDs in these populations.[1] All this can be achieved through increased resource allocation for NTDs and malaria healthcare provision; partnerships with local and international NGOs, and through community engagement to map and reach mobile populations.

Significant gaps persist in the current evidence base, highlighting critical needs for future research. None of the studies examined the role of socioeconomic factors in the occurrence of these diseases among migrants. Enhanced education, better healthcare access, and economic development, have been linked to substantial reductions in the burden of NTDs and malaria.[89] Further studies should specifically evaluate the impact of socio-economic factors on the burden of these condition in migrants in the MENA region. Moreover, we only found 11 out of 23 NTDs studies in three North African countries, and data regarding risk factors for NTDs in migrants were limited. Future research in the North African sub-region is critical as countries like Egypt are hosting millions of migrants from Sudan, Palestine, Syria, and Yemen as a result of regional conflicts, while countries like Morocco and Tunisia are transit destinations for migrants moving to Europe. However, such research is often challenging due to methodological and ethical challenges. A lack of trust in authorities or researchers and concerns related to privacy and informed consent together with high mobility of many migrants further contributes to low recruitment, limited participation, and poor long-term follow-up in studies involving migrants.[93] Undocumented migrants are also frequently excluded from studies due to legal and logistical constraints.

## Strengths and limitations

Our study has several limitations. First, although we searched for grey literature, we included only peer-reviewed articles because we did not find grey literature on the burden, risk factors, or clinical outcomes in migrants in the MENA region. Second, the available evidence was limited and highly fragmented across diseases, and outcomes –most studies did not differentiate between migrants groups, limiting the ability to estimate pool prevalence . We were unable to calculate pooled estimates for most diseases due to heterogeneity across studies, variations in disease definitions, diagnostic methods, migrant categories (i.e., refugees and undocumented migrants) and other outcomes, low data quality in some cases, and the limited number of studies. Although five studies reported similar outcomes for malaria, we did not pool their estimates because of differences in diagnostic methods and migrant categories. We were also unable to disaggregate analyses by year of arrival or assess publication bias. Third, data included in the meta-analysis exhibited high heterogeneity; consequently, we interpreted the results with caution.

## Conclusions

Migrants in multiple MENA countries are disproportionately affected by malaria and NTDs, highlighting their vulnerability. Future research should systematically assess the risk factors and clinical outcomes of malaria and NTDs among migrant populations particularly in North Africa to guide evidence-based approaches for effective prevention, early diagnosis, and treatment. [10,11]

## Author contributions

ARM conceptualised the idea for this project. FS planned the overall methodology for the review and co-led the write-up of this manuscript. EE and TM adapted the overarching method for each disease area and carried out all the processes for data collection and analyses; HM helped in the data analysis. AA, SE, FS, AH, MH, IB, and ARM supervised the review. All authors were involved in drafting and reviewing the manuscript. ARM is responsible for the overall content as guarantor.

## Public and patient involvement

Some members of the research team are migrants living in the MENA region and were involved in the design, conduct, and dissemination of the study.

## Data sharing

All data used in this review were previously published.

## Data Availability

All data used in this review were previously published.

## Acknowledgements

We would like to thank Sabina Gillsund and Narcisa Hannerz for developing the search strategies and all the members of The Middle East and North Africa Migrant Health Working Group: Adel Abdelkhalek (Badr University of Cairo, Adnene Ben Haj Aissa (Office National de la Famille et de la Population, Tunisia), Charles Agyemang (University of Amsterdam, Netherlands), Salma Altyib (Ministry of Health, Sudan), Ali Ardalan (WHO Regional Office for the Eastern Mediterranean), Hanen Ben Belgacem (IOM Tunisia), Imane Belkhammar (Maroc Solidarité Médico-Sociale MS2, Morocco), Thomas Calvot (Médecins du Monde, Tunisia), Nuria Casamitjana (University of Barcelona, Spain), Anissa Ouahchi (Office National de la Famille et de la Population, Tunisia), Hassan Edries Hasaan Mohammed (University of Gezira, Sudan), Oumnia Bouaddi (Mohammed VI University of Sciences and Health, Morocco), Moudrick Abdellatifi (Mohammed VI University of Sciences and Health, Morocco), Luciana Ceretti (IOM Morocco), Nelly Chavassieux (Maroc Solidarité Médico-Sociale MS2, Morocco), Hassan Chrifi (Ecole Nationale de Santé Publique, Morocco) Mohamed Douagi (Office National de la Famille et de la Population, Tunisia), Algdail Elnil (Sudan Organization Network for Peace & Development, Sudan), Gonzalo Fanjul (Institut for Global Health Barcelona, Spain), Fouad M Fouad (American University of Beirut, Lebanon), Chiaki Ito (IOM MENA), Abdedayem Khelifi (Ecole Nationale de Santé Publique, Morocco), Lora Makhlouf (Médecins du Monde, Morocco), Maissa Mokni (Médecins du Monde, Tunisia), Davide Olchini (Médecins du Monde MENA), Tarik Oufkir (Maroc Solidarité Médico-Sociale MS2, Morocco), Nasong Park (IOM Egypt), Shaima Abd Alrahman (IOM Egypt), Wessam El Nahry(IOM Egypt),Samir Hadjiabduli (IOM Egypt),Giuseppe Raffa (Médecins du Monde, Tunisia), Wafa Saidi (Ministry of Health, Tunisia), Sandra Santafé (Institut for Global Health Barcelona, Spain), Alice Sironi (IOM Tunisia), Fatma Temimi (Office National de la Famille et de la Population, Tunisia), Zeineb Turki (Médecins du Monde, Tunisia).

## Ethical considerations and consent

Patient consent for publication: Not applicable.

## Ethics approval

Ethical approval was not required to undertake the study because the study did not involve human participants.

## Annex 1. Search strategy

|  |  |  |
| --- | --- | --- |
| Interface: Ovid MEDLINE(R) and Epub Ahead of Print, In-Process & Other Non-Indexed Citations and Daily |  | Field labels<br>exp/ = exploded MeSH term<br>/ = non exploded MeSH term<br>.ti,ab,kf. = title, abstract and author keywords<br>adjx = within x words, regardless of order<br>* = truncation of word for alternate endings |
| Date of Search: February 27, 2023 |  |  |
| Number of hits: 692 |  |  |
| Comment: In Ovid, two or more words are automatically searched as phrases; i.e. no quotation marks are needed |  |  |
| Database(s): **Ovid MEDLINE(R) ALL** 1946 to February 24, 2023 |  |  |
| Search Strategy: |  |  |
| # | Searches | Results |
| 1 | exp Human Migration/ | 27722 |
| 2 | exp "Emigrants and Immigrants"/ | 15372 |
| 3 | "Transients and Migrants"/ | 14022 |
| 4 | Refugees/ | 12903 |
| 5 | Refugee Camps/ | 288 |
| 6 | (alien* or asile or asylum* or (border* adj2 cross*) or (countr* adj3 origin*) or diaspora or displace? or displacement* or emigrant* or emigration or expat? or expatriate? or foreigner* or foreign-born* or foreign background* or foreign population* or immigrant* or immigration or migrant* or migration or naturalized citizen* or new* arriv* or newcomer* or new-comer* or nomad* or non-citizen* or nonnative* or non-native* or nonnational or non-national or nonresident or non-resident* or resettlement* or re-settlement* or refugee* or settler* or squatter* or undocumented worker*).ti,ab,kf. | 573858 |
| 7 | or/1-6 | 587633 |
| 8 | exp Middle East/ | 159113 |
| 9 | exp Africa, Northern/ | 41057 |
| 10 | (Abu Dhabi or Ajman or Algeri* or Arab* or Bahrain* or Bahreiin* or Dubai or Egypt* or Emirat* or Fujairah or Gaza* or Golf* or Gulf* or Ifriqiya* or Irak* or Iraq* or Jorda* or Jumhuuriya* or Koweit* or Kuwait* or Kuwayt* or Leban* or Liban* or Liby* or Lubnan* or Maghr* or Maroc* or Maser* or Masr or Misr or MENA or Middle East* or Morocc* or North* Afric* or Oman* or Palestin* or Qatar* or Saudi* or Sharjah or Soudan* or Sudan* or Syria* or Syuri* or Tunis* or Uman* or Umm Al-Quwain or West Bank or Yemen*).ti,ab,kf. | 318043 |
| 11 | or/8-10 | 434382 |
| 12 | Neglected Diseases/ | 2424 |
| 13 | exp Malaria/ or exp Plasmodium/ | 90811 |
| 14 | Buruli Ulcer/ or Mycobacterium ulcerans/ | 1098 |
| 15 | exp Dengue/ or exp Arbovirus Infections/ or exp Flavivirus Infections/ | 52097 |
| 16 | Chikungunya Fever/ | 2723 |
| 17 | Dracunculiasis/ | 907 |
| 18 | exp Echinococcosis/ | 20591 |
| 19 | exp Trematode Infections/ | 37974 |
| 20 | Trypanosomiasis, African/ or Trypanosomiasis/ or Euglenozoa Infections/ | 10112 |
| 21 | exp Leishmaniasis/ | 25341 |
| 22 | exp Leprosy/ or Mycobacterium leprae/ | 24706 |
| 23 | Elephantiasis, Filarial/ or Wuchereria bancrofti/ | 4066 |
| 24 | Mycetoma/ or exp Dermatomycoses/ | 29882 |
| 25 | exp Onchocerciasis/ or exp Filariasis/ or Onchocerca volvulus/ | 16005 |
| 26 | Rabies/ or exp Rhabdoviridae Infections/ | 13177 |
| 27 | Scabies/ or exp Ectoparasitic Infestations/ | 22522 |
| 28 | exp Schistosomiasis/ | 24882 |
| 29 | exp Helminthiasis/ | 134192 |
| 30 | Snake Bites/ | 5405 |
| 31 | exp Taeniasis/ or exp Cestode Infections/ or exp Cysticercosis/ or Neurocysticercosis/ | 33945 |
| 32 | Trachoma/ or exp Conjunctivitis, Bacterial/ | 6181 |
| 33 | Treponemal Infections/ or Yaws/ or Treponema pallidum/ | 5990 |
| 34 | (neglected adj3 disease*).ti,ab,kf. | 8699 |
| 35 | ((arbovirus or echinococcus or euglenozoa or flavivirus or nematomorpha or rhabdoviridae or schistoma or trematode or treponemal) adj3 infection*).ti,ab,kf. | 3845 |
| 36 | (actinomycetoma or african sleeping sickness* or african trypanosomias?s or bacterial conjunctivitis or bancroftian elephantias?s or bancroftian filarias?s or bejel* or bilharsia* or bilharzias?s or black fever or borderline tuberculoid* or breakbone fever or break bone fever or buruli ulcer or chikungunya or coenuri* or coenuros?s or coenurus cerebralis or cysticercos?s or cysticercus cellulosae or deep mycos?s or dengue* or dermatomycos?s or dracuncul* or echinococcus?s or ectoparasitic infestation* or ectoparasitos?s or egyptian ophthalmia or eumycetoma or filarial elephantias?s or filariose or filiariasis or frambesia or guinea worm disease* or hansen* disease* or helminthias?s or hydatid cysts or hydatidos?s or hydrophobia or kalazar or kala azar or katayama fever or leishmani* or leprae or leepre or leprosie or leprosy or lymphatic filarias?s or lyssa or lyssas or madura foot or maduromycosis or malare or malaria or malayi elephantias?s or malayi filarias?s or mycetome or mycetoma or mycobacterium ulcerans or nagana or neurocysticercos?s or neuroschistosomias?s or onchocercias?s or onchocerca volvulus or onchocercose or oriental sore or plasmodium falciparum or paludisme or rabies or raby or river blindness or sarcoptic mange or scabies or schistosomias?s or schistosomal myel* or snakebite* or snake bite* or snake envenoming or taenia* or taenias?s or teeniase or trachoma* or treponema pallidum or treponematos?s or trypanosomiase or trypanosoma or trypanosomiasis or ulceere or ulcer disease* or wuchereria bancrofti or yaw or yaws or zazzabin cizon sauro).ti,ab,kf. | 350022 |
| 37 | or/12-36 | 543044 |
| 38 | 7 and 11 and 37 | 1064 |
| 39 | (animals not humans).sh. | 5062487 |
| 40 | 38 not 39 | 967 |
| 41 | limit 40 to yr="2000 -Current" | 692 |

## Annex 2: Variables reported in the data extraction forms

### Study details

1. First author surname or name of organisation
2. Year of publication
3. Country
4. Number of centres
5. Study design
6. Study setting
7. Total study duration
8. Funding
9. Aim of study
10. Disease (for those who have multiple diseases under consideration)

### Methods details

11. Recruitment dates
12. General definition of sample
13. Participant inclusion criteria
14. Participant exclusion criteria
15. Definition of participants and comparator groups
16. Sample size at baseline for each participant group and total in the study
17. Sample size analysed for each participant group and total in the study
18. Numbers lost to follow-up/withdrawals for each participant group and total in the study
19. Follow-up duration (where appropriate)
20. Intervention and details for this
21. Baseline characteristics for each participant group and total in the study
  a. Age (as reported: mean, median, range, standard deviation, or frequencies and % in age categories)
  b. Female (frequency, %)
  c. Country of birth (frequency, %)
  d. Migrant type for migrant groups (all migrants, refugees, asylum seekers, labour migrants, internally displaced people, etc. and frequency/% of each as appropriate)
  e. Time in host country for migrant groups (as reported: mean, median, range, standard deviation, or frequencies and % if in categories)
  f. Living situation (frequency/% in camp setting, community, etc.)
  g. Occupation (frequency/%/% of each)
  h. Socioeconomic status (as reported)
  i. Co-morbidities
  j. Treatments
  k. Others

22. Outcomes and the definitions for each outcome

### Results by each outcome

*Indicators (e.g., incidence, mortality, coverage, uptake)*

23. Number/mean reported with outcome in each participant and comparator group
24. Number/mean reported without outcome in each participant and comparator group
25. Totals / mean in each group and with and without outcome
26. Statistical parameter reported, e.g., odds ratio, risk ratio, mean difference and 95% CI and/or p value
Crude (no adjustment) result reported
Adjusted result reported
Covariates adjusted for

(All the above in the results section should be repeated for each outcome of relevance reported in a study, as necessary)

#### Facilitators and barriers

27. List of barriers
28. List of facilitators
29. Other

### Conclusions

1. 30. Author’s conclusions
2. 31. Reviewer’s notes

## Annex 3. General characteristics included studies on neglected tropical diseases and malaria

| Study | Study period | Study country | Study setting | Diagnostic method | Study population | Sample size | Age inclusion criteria | Female (%) | Outcome(s) |
| --- | --- | --- | --- | --- | --- | --- | --- | --- | --- |
| Neglected Tropical Diseases |  |  |  |  |  |  |  |  |  |
| Soil-transmitted helminthic infection |  |  |  |  |  |  |  |  |  |
| Abu-Madi[53]<br>2010 | 2005-2008 | Qatar | HMC hospitals | Microscopic stool examination | Patients attending the hospital | Long-term migrants: 5898<br>Qatari citizens: 3310 | Any | 3881 (42.1) | Prevalence<br>Risk factors |
| Abu-Madi[57]<br>2011 | 2008 | Qatar | HMC medical commission | Microscopic stool examination | Screened cases | Newly arrived migrants: 766<br>Resident migrant workers: 772 | 18-52 | 532(34.5) | Prevalence<br>Risk factors |
| Abu-Madi[54]<br>2013 | 2005-2011 | Qatar | HMC hospitals | Microscopic stool examination | Screened cases | Long-term migrants: 12,150<br>Qatari: 6413 | Any | 7772 (41.8) | Prevalence<br>Risk factors |
| Abu-Madi[55]<br>2016 | 2005-2014 | Qatar | HMC hospitals | Microscopic stool examination | Screened cases | Long-term migrants: 19,929<br>Qatari 9357 | Any | 12,295(41.9) | Prevalence<br>Risk factors |
| Abu-Madi[56]<br>2016 | 2012-2014 | Qatar | Medical commission | Microscopic stool examination | Screened cases | Newly arrived migrant: 2486 | 16-58 | 1351 (54.3) | Prevalence<br>Risk factors |
| Dafalla[58]<br>2017 | Jan-Dec 2013 | UAE | Municipality public health clinic | Microscopic stool examination | Screened cases | Migrants: 21,347 | ≥16 | 3202 (15.0) | Prevalence |
| Imam[59]<br>2015 | Two months | Saudi Arabi | Medical check-up & report center | Microscopic stool examination | Screened cases | Non-nationals migrant: 120 | 20-55 | 63(52.5) | Prevalence |
| Schistosomiasis |  |  |  |  |  |  |  |  |  |
| Abu-Madi[57]<br>2011 | 2008 | Qatar | HMC medical commission | Microscopic stool examination | Screened cases | Newly arrived migrants: 766<br>Resident workers: 772 | 18-52 | 532 (34.5) | Prevalence |
| Al-Awadhi[66]<br>2021 | Jan 2010<br>Dec 2018 | Kuwait | Reference national laboratory | Serological screening and Microscopic stool examination | Suspected cases | Migrants: 916<br>Kuwaiti citizens: 61 | 1-84 | 474 (47.8) | Confirmed cases (%)<br>Risk factors |
| Balahbib[68]<br>2020 | 2005-2017 | Morocco | Epidemiology directorate | NR | Reported cases | Migrants: 27 | 8-52 | 2 (7.4) | Distribution of cases by migration |
| Imam[59]<br>2015 | Two months | Saudi Arabia | Medical check-up & report center | Microscopic stool examination | Screened cases | Migrants: 120 | 63 (52.5) | 63 (52.5) | Prevalence |
| Mohammed[67]<br>2014 | Jan 2012-Dec 2013 | Saudi Arabia | Albaha PHC centers | Microscopic stool examination | Suspected cases | Migrants: 1891<br>Saudi nationals 232,364 | Any | 125,445(54.0) | Confirmed cases (%) |
| Teniasis |  |  |  |  |  |  |  |  |  |
| Abu-Madi[57]<br>2011 | 2008 | Qatar | HMC Medical Commission | Microscopic stool examination | Screened cases | Newly arrived. migrant 766;<br>Resident workers 772 | 18-52years | 532 (34.5) | Prevalence |
| Abu-Madi[56]<br>2016 | 2012–2014 | Qatar | Medical Commission | Microscopic stool examination | Screened cases | Newly arrived migrants 2486 | 16-58 year | 1351 (54.3) | Prevalence |
| Al-Awadhi[77]<br>2019 | 2015 -2017 | Kuwait | Community-based | Stool Microscopic, Blood: ELISA | Screened | Migrant 1000 | 21 to 65 years | 500 (50) | Prevalence<br>Risk factors |
| Dafalla[58]<br>2017 | Jan – Dec 2013 | United Arab Emirates | Sharjah Municipality Microscopic Public Health Clinic | Microscopic stool examination | Screened cases | Migrants 21,347 | ≥ 16 years | 3,202 (15) | Prevalence |
| Haddad[78]<br>2023 | 2013-2018 | Qatar | Hamad Medical Corporation | Radiographic image, histology EEG | Reported cases | Migrants 415 (98.8)<br>National 5 (1.2) | Any age | 27 (6.4) | Confirmed cases of neurocysticercosis (%) |
| Hassan[60]<br>2020 | Aug 2016-May 2017 | Iraq | Refugee camp | Microscopic stool examination | Suspected cases | IDPs 3925 | 8month -60 year | NR | Confirmed cases of taenia (%) |
| Imam[59]<br>2015 | Two months | Saudi Arabia | Medical Check-up & Report Center | Microscopic stool examination | Screened cases | Migrants 120 | 20-55 year | 63 (52.5) | Prevalence |
| Cystic echinococcosis |  |  |  |  |  |  |  |  |  |
| Al-Awadhi[66]<br>2021 | Jan 2010 -<br>Dec 2018 | Kuwait | Ref. national laboratory | Serological screening | Suspected cases | Migrant 916<br>Kuwaiti 61 | 1-84 years | 474 (47.8) | Confirmed cases (%)<br>Risk factor |
| Filariasis |  |  |  |  |  |  |  |  |  |
| Iqbal[79]<br>2006 | Apr 2000 -<br>Nov 2003 | Kuwait | Ports and borders center, Ministry of Health | NFB microscopy<br>ICT<br>TropBio ELISA assay. | Screened cases | Migrant worker 1050 | 14-61 years | 447 (42.6) | Prevalence |
| Cutaneous leishmaniasis (CL) |  |  |  |  |  |  |  |  |  |
| Cross-sectional study |  |  |  |  |  |  |  |  |  |
| Abdulla[46]<br>2018 | January -<br>December 2016 | Iraq | Ebril Preventive health directorate | Clinical | Reported cases | IDPs: 49 (20.9)<br>Refugees: 18(7.7)<br>Residents: 167 (71.4) | Any age | 83 (35.5) | Distribution of migrants among confirmed cutaneous leishmaniasis |
| Abdalla[42]<br>2019 | Mar2010-Sep2011 | Saudi Arabia | Almadina Surveillance system | Microscopically | Reported cases | Non-national: 132 (80.5)<br>National: 32 (19.5) | Any age | 0 | Distribution of migrants among confirmed cutaneous leishmaniasis |
| Alawieh[47]<br>2014 | Jan 2001 -<br>Mar 2014 | Lebanon | Surveillance system | Positive smear or culture. | Reported cases | Syrian refugee: 1206 (96.5)<br>Nationals / Palestinian refugees: 44 (3.5) | Any age | 527 (51) | Distribution of migrants among confirmed cutaneous leishmaniasis |
| Alhawarat[44]<br>2020<br>Amr[45]<br>2018 | 2010-2016<br>(2012-2016) <sup>1</sup> | Jordon | Surveillance system | Positive smear or culture | Reported cases | Syrian refugee: 559 (45.0)<br>Nationals: 646 (52.0) | Any age | 489 (39.3) | Distribution of cases by migration |
| Alraey[43]<br>2022 | Jan - Dec 2021 | Saudi Arabia | Asir Surveillance system | Microscopic PCR | Reported cases | Non-national: 18 (9.3); National: 176 (90.7) | Any age | 73 (37.6) | Distribution of migrants among confirmed cutaneous leishmaniasis |
| Amin[41]<br>2013 | 2000-2010 | Saudi Arabia | Al-Ahsa city, Surveillance system | NR | Reported cases | Non-national: 5393(54.1)<br>National: 4569 (45.9) | 3 months to 81 years | 2172 (21.8) | Distribution of migrants among confirmed cutaneous leishmaniasis |
| Farah[50]<br>2023 | 2013-2019 | Lebanon | Communicable disease surveillance Systems | NR | Confirmed cases | Syrian refugee 1683 | Any age | NR | Incidence |
| Hassanein[51]<br>2022 | January 2006 to December 2021 | Saudi Arabia | Vector-borne Diseases Control Unit Tabuk | Microscopic | Reported cases | Migrant 739(46.9)<br>Saudi national 836(53.1) | Any age | 265(17) | Distribution of migrants among confirmed cutaneous leishmaniasis |
| Saroufim[48]<br>2014 | Sep 2012 | Lebanon | Refugee camp | Microscopic PCR | Suspected cases | Refugee 948 | Any age | 527 (51) | Confirmed cases (%) |
| Suliman[49]<br>2019 | May 2013 | Sudan | Village | Tissue smear Microscopy | Screened | IDPs 1236 | Any age | 688 (55.7) | Prevalence |
| Case-control study |  |  |  |  |  |  |  |  |  |
| Lehlewa[52]<br>2021 | NR | Iraq | Districts in Diyala Province (Al-Muqdadiya and Al-Mansuriya) | Clinical | Cases of CL and Control | Cases 432 (51%)<br>Control 415(49%) | Any age | 407 (48.2) | Risk factors |
| Dengue |  |  |  |  |  |  |  |  |  |
| Ahmed[64]<br>2022 | Aug 2015 - Feb 2016 | Sudan | Refugee camp | RT-PCR | Suspected cases | IDPs 204 | Any age | NR | Confirmed cases (%)<br>Clinical outcomes |
| Alqassim[65]<br>2024 | Jan 2015 Aug 2020 | Saudi Arabia | Department of Health surveillance system- Jizan | PCR Serology | Reported cases | Migrant 910(26.6)<br>Saudi 2517(73.4) | 0-95 year | 94 (35.2) | Incidence |
| Alshabi[62] 2019 | 2012-2020 | Saudi Arabia | Hospital-based | Serologically PCR | Suspected cases | Migrant 1752<br>Saudi 4885 | 0-100 year | 2471 (37.2) | Confirmed cases (%) |
| Farag[63] 2022 | Jan 2013 - Dec 2021 | Qatar | Surveillance & outbreak unit | PCR | Reported cases | Migrants 163(98)<br>Qatari 2(2) | Any age | 140 (24) | Reported cases |
| Humphrey[61] 2019 | Jan 2013 - June 2016 | Qatar | HMC (Blood bank) | ELISA | Screened cases | Migrants 1792<br>Qatari 200 | >18 years | 0 | Prevalence<br>Risk factors |
| Chikungunya |  |  |  |  |  |  |  |  |  |
| Humphrey[61] 2019 | Jan 2013 - Jun 2016 | Qatar | HMC (Blood bank) | ELISA | Screened cases | Migrants 1792<br>Qatari 200 | >18 years | 0 | Prevalence<br>Risk factors |
| Scabies |  |  |  |  |  |  |  |  |  |
| Abdullah[73] 2020 | Jan -Aug 2019 | Iraq | Dhokuk province health centre-IDP camp | Clinical Microscopically Dreamscape | Suspected cases | IDP 6264<br>residents 29,183 | Any age | NR | Confirmed cases (%) |
| ALBERFKANI[74] 2020 | Jan 2018 - Dec 2019 | Iraq | Cham Mishiko city IDP camp | Microscope | Suspected cases | IDP 1300 | >=1year | 654 (50.30) | Confirmed cases (%) |
| Alsamara[72] 2017 | Oct 2015 - Oct 2016 | Iraq | Kirkuk city Dermatologic clinic | Clinical Microscopic | Reported cases | IDP 153 | Any age | 68 (44.4) | Distribution of cases by migration |
| Trachoma |  |  |  |  |  |  |  |  |  |
| Javaloy[71] 2002 | Oct 2001 | Algeria | Refugee camp | Clinical examination | Screened cases | children aged 3-16) 527 | 3-16 year | 274 (52) | Prevalence |
| Macleod[70] 2019 | 2014 - 2015 | Sudan | IDPs Camps | clinical examination | Screened cases | IDPs Children 1823;<br>adult 2139 | Any age | 2548 (56) | Prevalence |
| Sanders[69] 2019 | NR | Sudan | Refugee camp | Clinical examination | Screened cases | South Sudanese refugee: Children 1601; Adult 1734 | Any age | 1986 (59.5) | Prevalence |
| Leprosy |  |  |  |  |  |  |  |  |  |
| Al-Mutairi[75] 2010 | Jan 2003 - Dec 2008 | Kuwait | Farwaniya dermatology departments | The detection of lepra bacilli slit skin smear. nasal smears | Reported cases | Migrants 41(89.1)<br>Kuwaiti 5(10.9) | Any age | 8 (17.4) | Distribution of cases by migration |
| Assiri[76]<br>2014 | 2003 - 2010 | Saudi Arabia | Surveillance system | WHO criteria | Reported cases | Migrants 39(57.4)<br>Saudi 139(42.6) | 15-44 | Total 103(42.5) | Distribution of cases by migration |
| Malaria |  |  |  |  |  |  |  |  |  |
| Abdalal[28]<br>2023 | 2015-2018 | Saudi Arabia | Jizan Aledabi and Baish centre | Malaria rapid diagnostic test or microscopy | Reported cases | Non-Saudi 2407<br>Saudi: 803<br>Cross border | IQR 25 | 324 (10) | Distribution of cases by migration<br>Risk factors |
| AlKuwari[27]<br>2009 | 2004 - 2006. | Qatar | Surveillance system | NR | Reported cases | Migrant 434(99.1)<br>Qatari 4 (0.9) | Any age | 70(16) | Distribution of cases by migration<br>Risk factors |
| AlRumhi[25]<br>2020 | 2013 - 2016 | Qatar | HMC hospital | Microscopic | Suspected | Migrants 583 | NR | NR | Confirmed cases (%)<br>Risk factors |
| Anderson[36]<br>2011 | Jan 2006 - Dec 2009 | Sudan | UNHCR database | RDT or Microscopy | Reported cases | NR | Any age | NR | Clinical outcomes |
| Elguarisha[29]<br>2019 | Sep 2014- Nov 2017 | Sudan | Hospitals and health centers in 10 states | Microscopy | Suspected cases | Refugees: 384<br>Residents: 842 | 0-85 | 40 | Confirmed cases (%)<br>Risk factors |
| EL-MALKY[37]<br>2016 | 2014 -2015 | Saudi Arabia | Centre of Malaria Elimination program | Microscopy | Reported cases | Migrant 232 (98.7)<br>Saudi 3 (1.3) | >= year | 89(38) | Distribution cases by migration<br>Risk factors |
| ElMekki[30]<br>2012 | Nov 2008 Feb 2009 | Sudan | Khartoum Refugee camp | Microscopic | Suspected | IDPs 300 | Any age | NR | Confirmed cases (%)<br>Clinical outcomes |
| Eshaq[38]<br>2020 | Jul 2018 - Dec 2018 | Sudan | AlGeneina Ardamata IDP camp. | nPCR | Suspected | IDPs 365<br>Visitor 15 | 1 to 80 years | 222(58.4) | Confirmed cases (%) |
| Farag[32]<br>2018 | Jan 2008 - Dec 2015 | Qatar | Surveillance System | Microscopic | Suspected | Migrants 4078 (99.6);<br>Qatari 14 (0.4) | Any age | 284 (7) | Distribution of cases by migration |
| Hawash[35]<br>2019 | Aug 2016-Sep 2018 | Saudia Arabia | Tertiary care hospital | PCR | Suspected | Migrants 134<br>Saudi 141 | >= 5 years | 165 (60) | Confirmed cases (%)<br>Risk factors |
| Iqbal[26]<br>2020 | Jan 2013 – Dec 2018 | Kuwait | Infectious Diseases Hospital | Rapid diagnostic test | Reported cases | Migrant 1895(99)<br>Kuwaiti 18 (1) | 1 to 73 years | NR | Distribution of cases by migration |
| Khan[39] 2006 | 2000-2002 | Qatar | HMC Hospital | Microscopic | Reported cases | Migrant 147 (96)<br>Qatari 6 (4) | Any age | 35(22.9) | Distribution of cases by migration<br>Risk factors |
| Khan[34] 2009 | Jan 2005 – Jun 2005 | Qatar | HMC hospital | Microscopy | Reported cases | Migrant 77 (95)<br>Qatari 4 (5) | Any age | 17(21) | Distribution of cases by migration<br>Clinical outcomes |
| Memish[40] 2014 | 2008 -2011 | Saudi Arabia | Makkah: 19 sentinel sites | NR | Reported cases | Migrant 302(95)<br>Saudi 16(5) | Any age | NR | Distribution cases by migration |
| Nilles[33] 2014 | Jan 2008 - Dec 2010 | Dubai Emirate | 599-bed Rashid Hospital and Trauma Center | NR | Reported cases | Migrant 629(100) | Any age | 44(7) | Distribution of cases by migration<br>Risk factors<br>Clinical outcomes |
| Saeed[31] 2003 | No 2001 – Dec 2002. | Sudan | Khartoum IDPs camp | NR | Suspected | IDPs 856 | Any age | 734(85.7) | Incidence<br>Risk factors |

Note: All studies are cross-sectional unless stated otherwise in the table. <sup>1</sup> Two studies in the same population. Abbreviations: HMC: Hamad Medical Corporation, UAE: United Arab Emirates, NR: Non-reported, PCR: polymerase chain reaction, nPCR: Nested polymerase chain reaction, IDP: Internally displaced persons, ICT: Immunochromatographic Test, ELISA: Enzyme-linked Immunosorbent Assay, UNHCR: United Nations High Commissioner for Refugees, RT-PCR: Reverse Transcriptase Polymerase Chain Reaction

## Annex 4. Risk factors for malaria and NTDs among migrants in the Middle East and North Africa

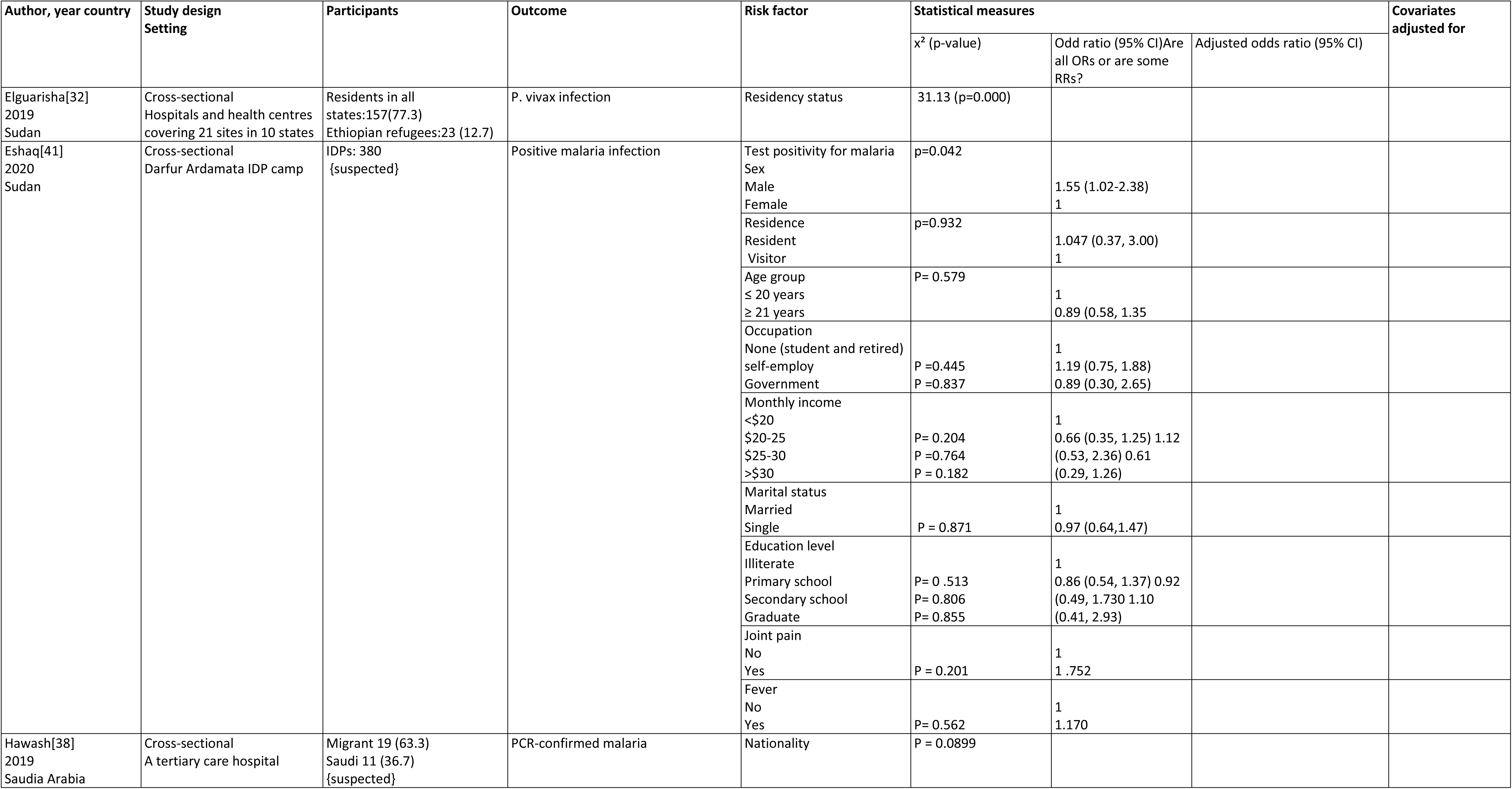
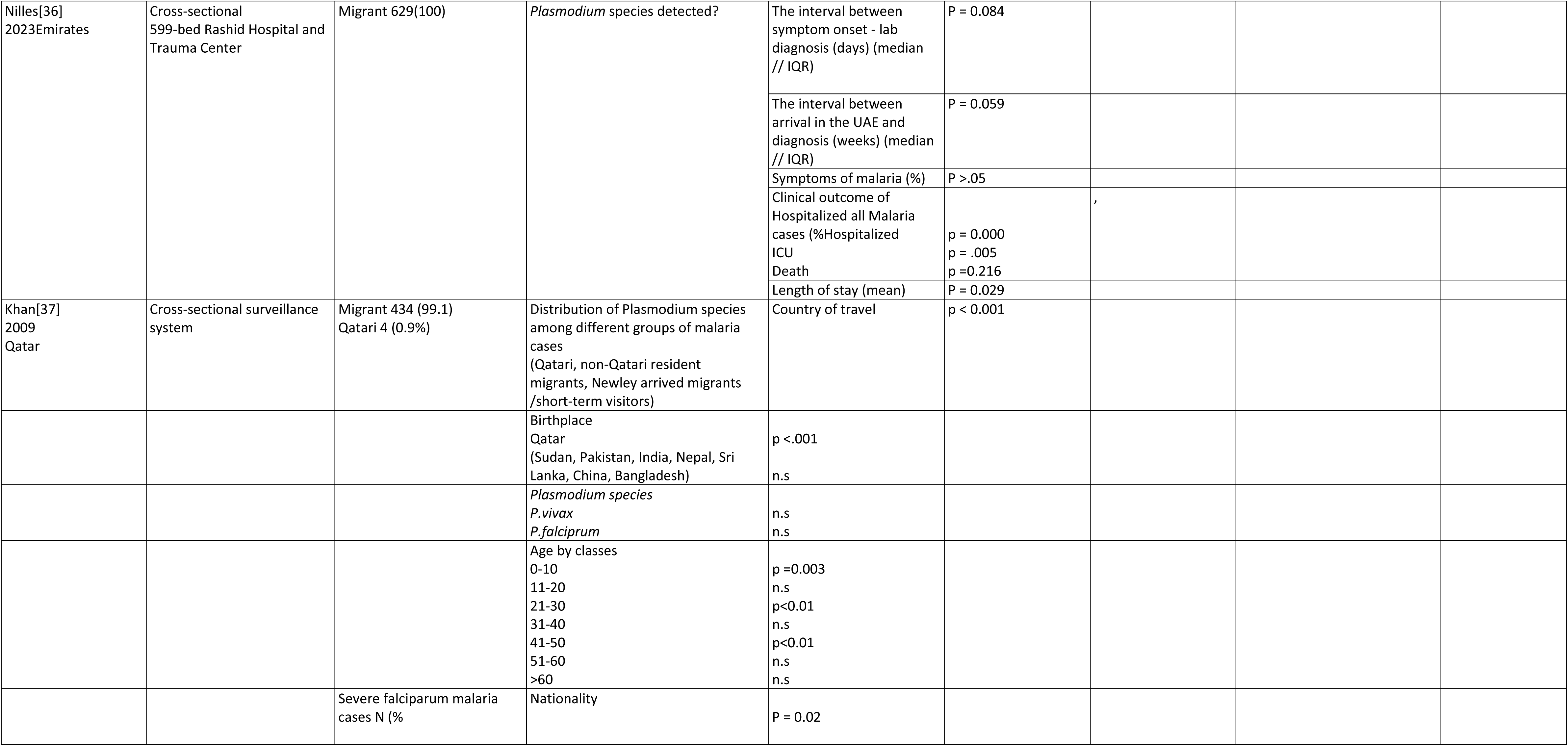
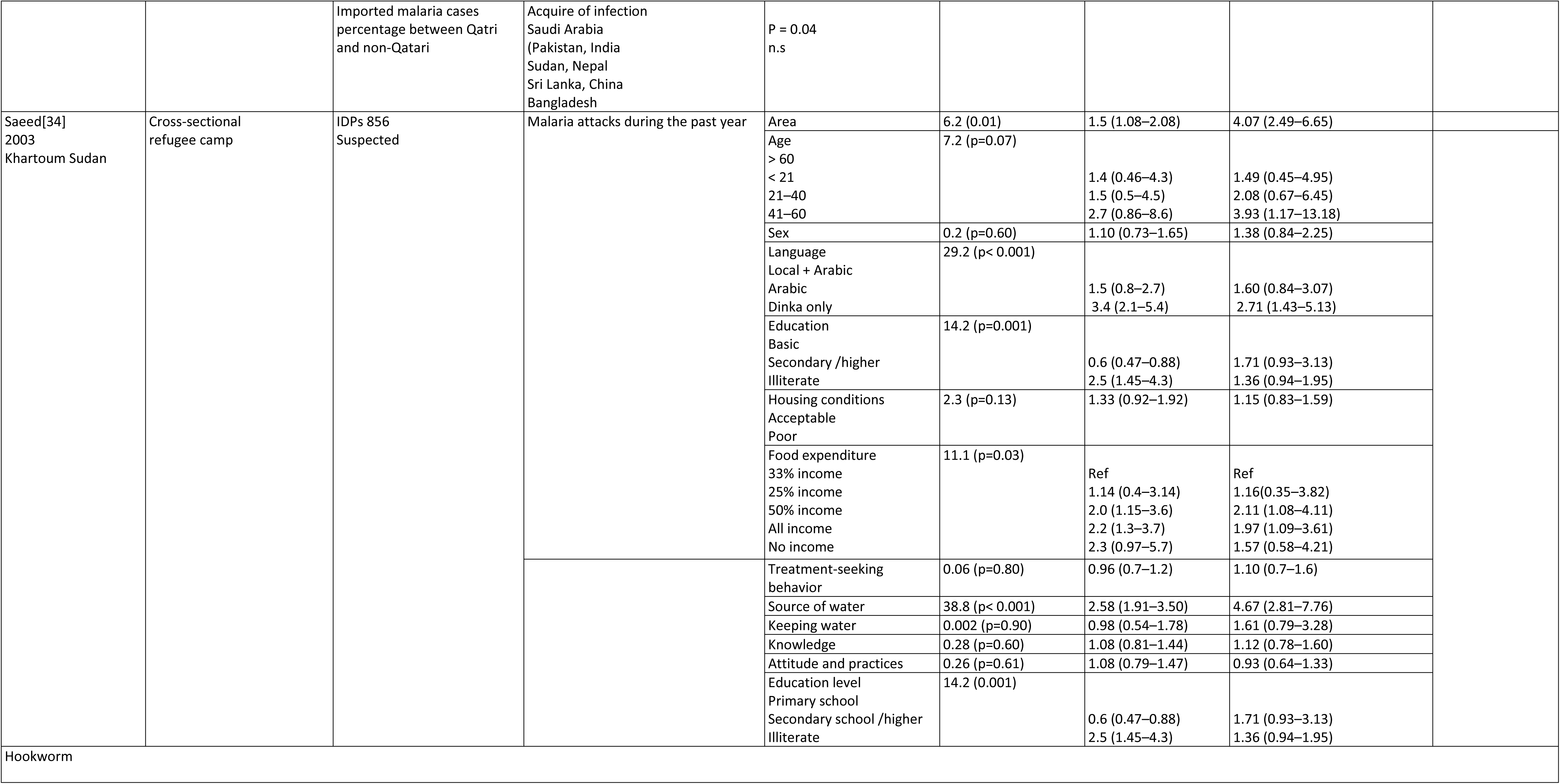
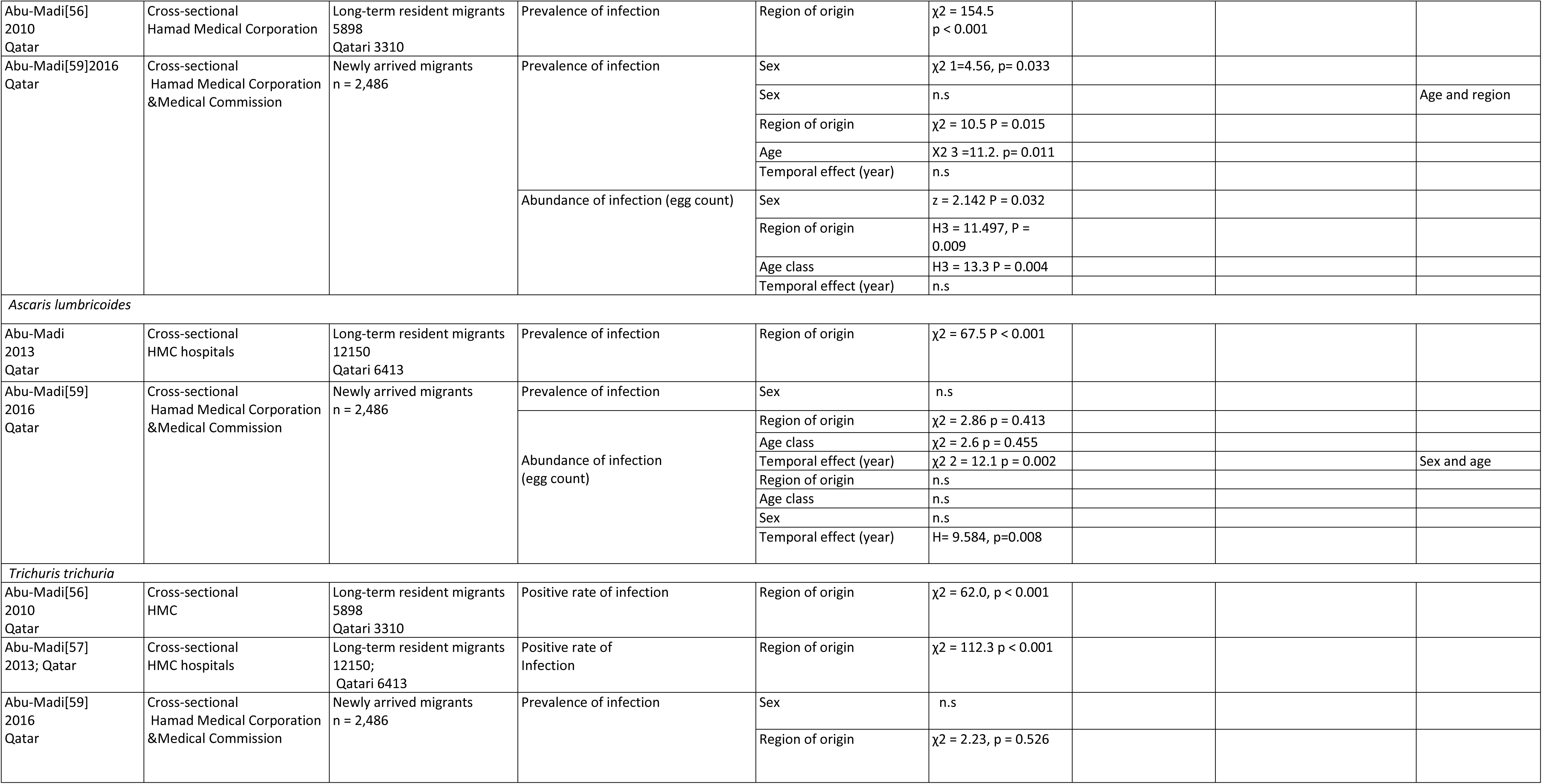
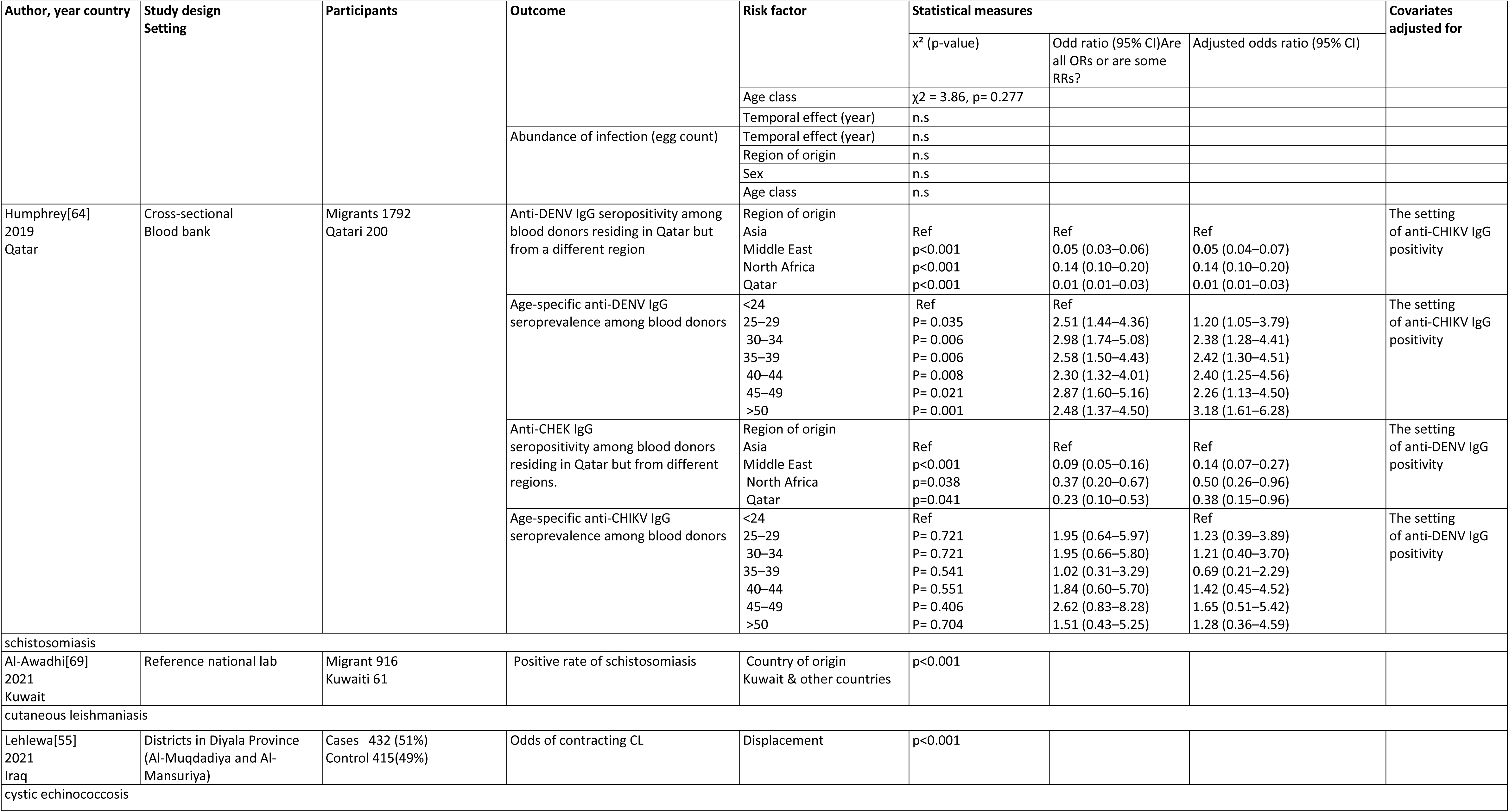
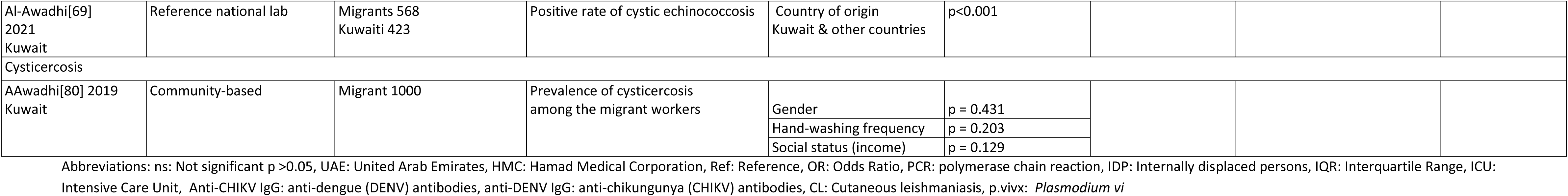

## Annex 5. Distribution of cutaneous leishmaniasis cases among migrants and non-migrants in the MENA region

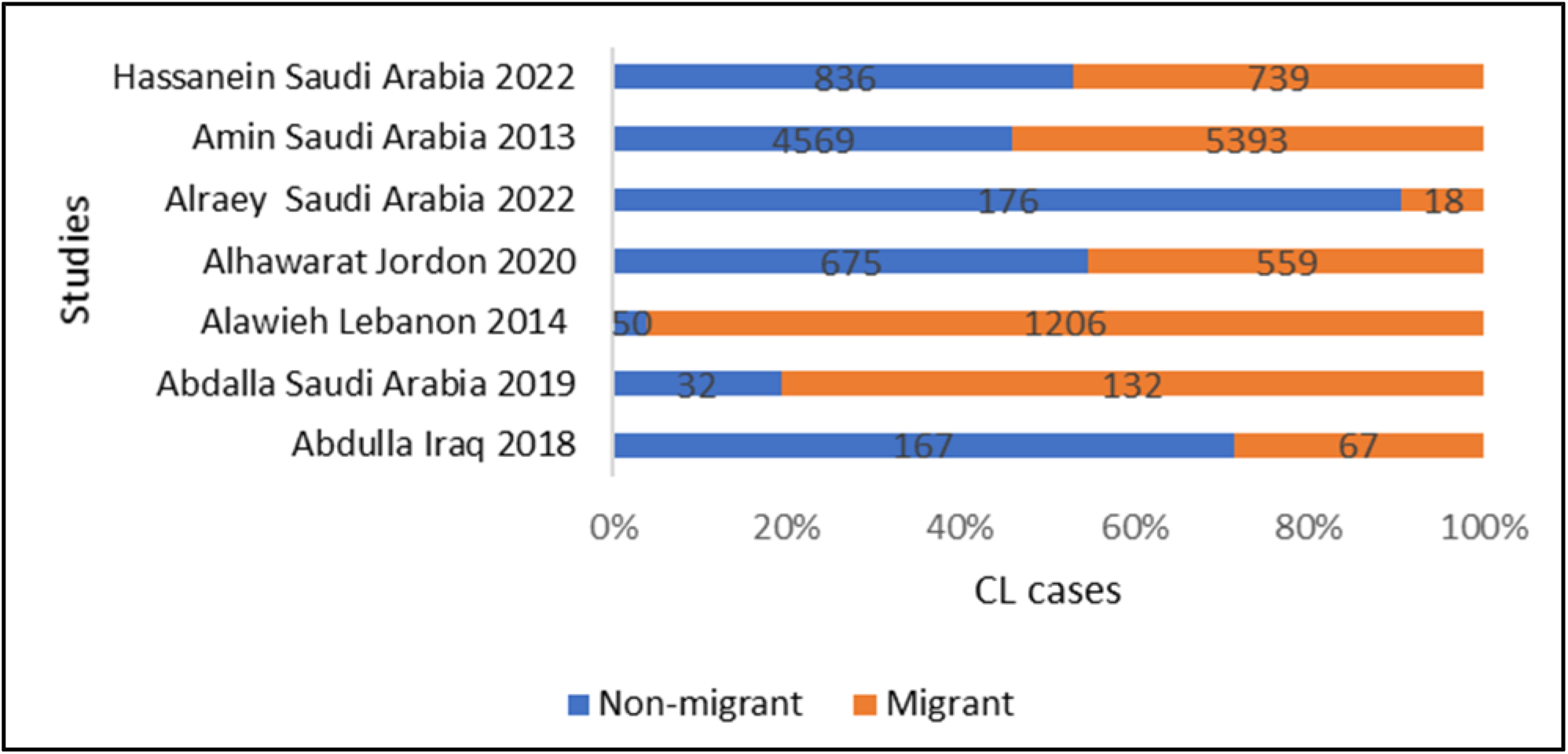
Abbreviation: CL: Cutaneous leishmaniasis

## Annex 6. Distribution of malaria cases among migrants and non-migrants in the MENA region

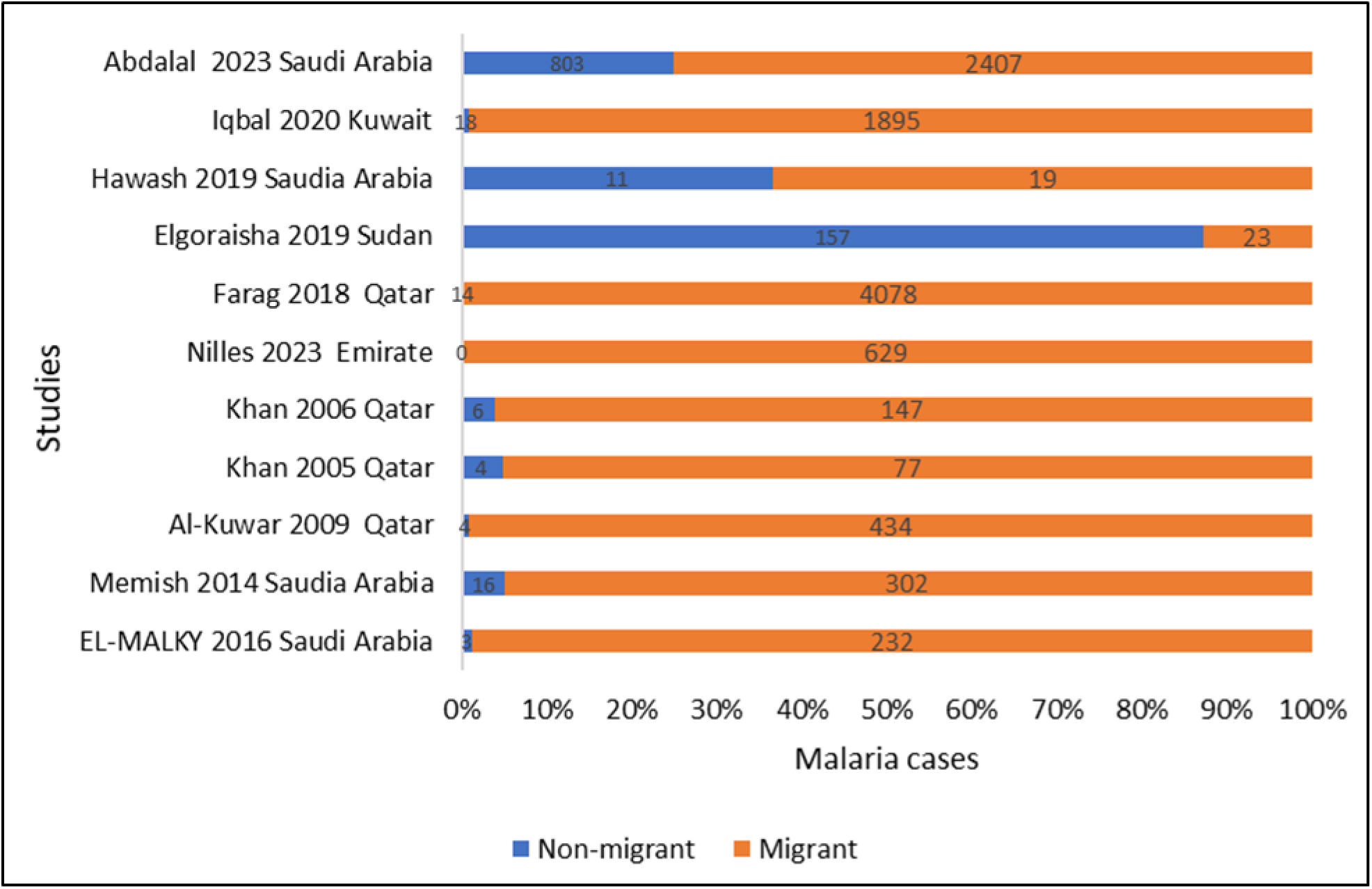

